# PGS Browser: a public platform for personalized polygenic score interpretation

**DOI:** 10.1101/2025.07.03.25330851

**Authors:** Nikita Kolosov, Mary P. Reeve, Pietro Della Briotta Parolo, Mitja I. Kurki, Vincent Llorens, FinnGen, Timo Petteri Sipila, Adam Herman, Ivan Molotkov, Mervi Aavikko, Samuli Ripatti, Aarno Palotie, Mark J. Daly, Mykyta Artomov

## Abstract

Identifying individuals at elevated risk before disease onset is a cornerstone of personalized prevention, and polygenic scores (PGSs) have proven to be instrumental in this regard. In this study, we systematically benchmarked 3,168 PGS models from the PGS Catalog in 473,681 FinnGen participants, revealing top-performing scores across major ancestry groups and highlighting traits where non-target PGSs significantly improve prediction. We conducted 10,531 phenome-wide association studies, uncovering 439,070 significant associations between PGSs and 4,739 clinical endpoints. To support individual score interpretation, we provide public access to ancestry-adjusted reference PGS distributions derived from the large-scale FinnGen cohort, alongside elastic-net Cox models for predicting time-to-event for 22 major disorders All data and predictive tools are accessible via the PGS Browser (pgs.nchigm.org), an interactive web-based platform for PGS analysis and interpretation. We envision that this esource will greatly advance both research and translational applications of polygenic scores.

## Introduction

Polygenic scores (PGSs) aggregate the effects of multiple genetic variants, derived from independent genome-wide association studies (GWAS), to estimate individual inherited susceptibility to complex traits. They have shown substantial utility for risk stratification, disease prediction, and exploration of shared genetic etiology across phenotypes through phenome-wide association studies (PheWAS)^1–6^

The Polygenic Score Catalog^7^ ^8^ is a centralized database storing 3,688 PGS models across 1,450 traits. The database also provides details on the origin of each PGS model and contributor-supplied validation metrics. The China Medical University Hospital cohort has recently been used to perform an external evaluation of PGS Catalog models, but, because most existing PGSs were developed in European cohorts, assessments using East Asian datasets may not be fully informative^9^ Without standardized benchmarking, cross-model comparisons are challenging, and the extent to which any given PGS could be applied to a new cohort remains unclear, thereby limiting clinical application.

Another significant barrier to bringing PGS into routine clinical practice is the need for reference PGS distribution to interpret an individual patient’s scores^10^ PGS distributions derived from major biobanks could be suitable for this purpose however, data-sharing regulations usually prohibit open public access to PGS data. Therefore, most efforts in interpreting individual scores are based on reference distributions from small public datasets, such as the 1000 Genomes^11^ and Human Genome Diversity Project^12^ The limited size of such reference datasets strongly underscores the need for secure resources that provide aggregated, ancestry-adjusted PGS distributions from large cohorts to facilitate and standardize the interpretation of PGS values.

In this study, address these challenges by integrating 3,168 PGS models, primarily sourced from the PGS Catalog, with extensive clinical data from 473,681 participants in the FinnGen cohort^13^ For every score we computed a comprehensive set of performance metrics and conducted a PheWAS across 4,739 disease endpoints, identifying a total of 439,070 experiment-wide significant associations. We also revealed diseases where a combination of scores yielded a increase in predictive performance and developed multivariate models that combine a PGS with age and sex to predict time-to-event for 22 major diseases. To facilitate broad adoption and access, generated ancestry-adjusted reference distributions for all 3,168 scores and developed the PGS Browser a interactive web interface paired with a command-line tool. The platform offers a single point of entry for FinnGen-based validation metrics, PGS-based PheWASs, ancestry-adjusted reference distributions, and predictive models, thereby moving PGSs from academic research toward practice-oriented, clinical implementation.

## Results

### PGS Catalog and FinnGen data harmonization

We evaluated polygenic score (PGS) models from the PGS Catalog using the FinnGen cohort as a independent validation dataset (**Fig. 1a**). FinnGen provides genetic and phenotypic data for 473,681 ndividuals (Release R11), comprising approximately 10% of Finland’s population and including 19,947 individuals of non-Finnish ancestry^13^ Its large cohort size, detailed longitudinal records, and minimal overlap with the development samples used to construct most PGS models (in contrast to the substantial overlap in the UK Biobank; **Supplementary Fig. 1a-b**) make it a suitable resource for external validation of PGS Catalog models.

**Figure 1.**
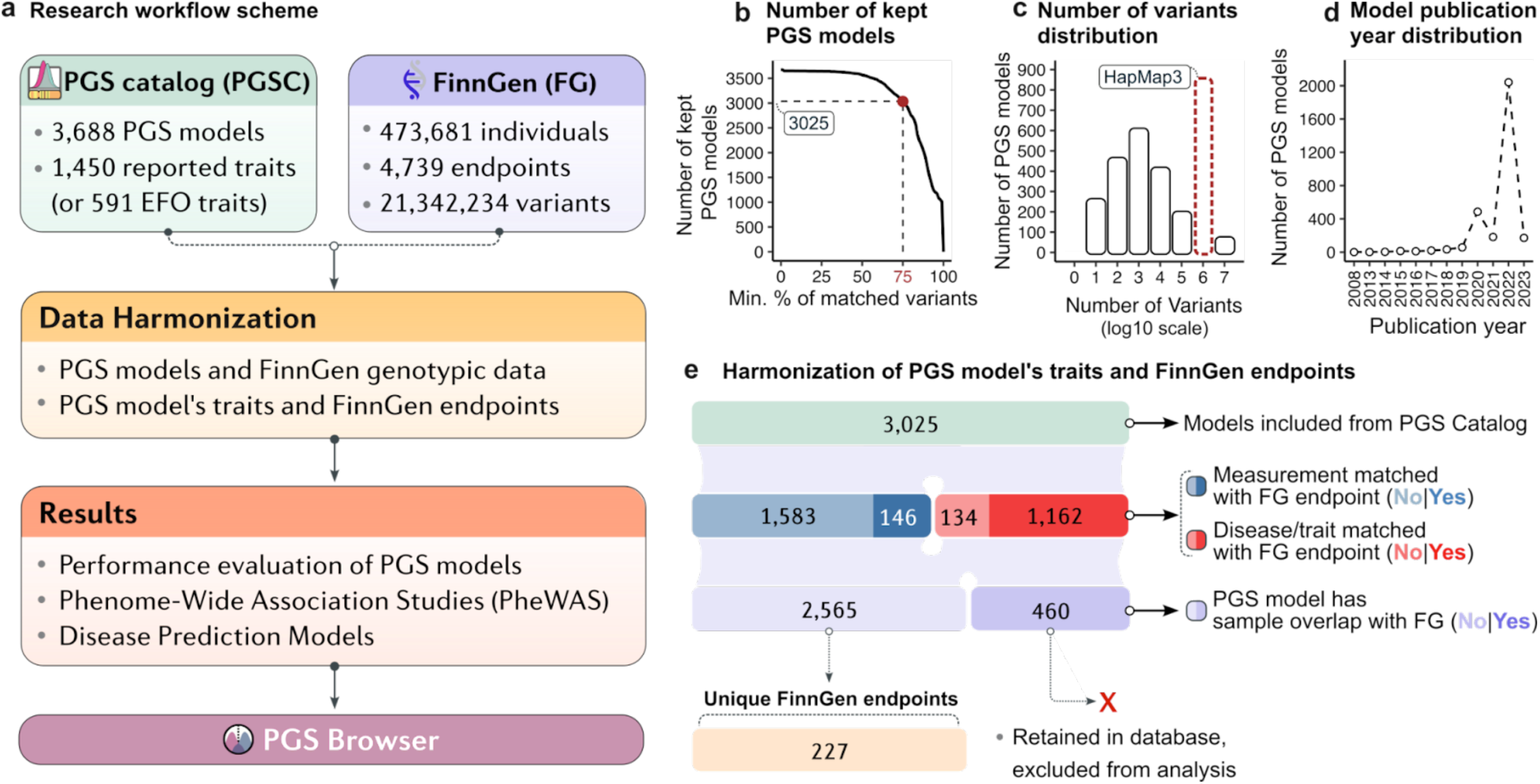
Harmonization of PGS Catalog and FinnGen data. (a) Overview of the article workflow, illustrating the harmonization of PGS Catalog and FinnGen data, subsequent experiments, and the development of the PGS Browser. (b) Total number of kept PGS models, based on the threshold for the percentage of matched variants. (c) Distribution of the number of variants in PGS models (log10 scale), with the brown column mainly consisting of models that used HapMap3 variants. (d) Number of PGS models by the year of publication used for their derivation. (e) Process of harmonizing PGS model traits with FinnGen endpoints. Opaque red and blue colors represent matched PGS models, while transparent red and blue colors indicate PGS models without corresponding FinnGen endpoints. Light purple represents models with sample overlap between the cohort used for PGS model development and FinnGen cohorts. Opaque purple indicates models with sample overlap, and transparent purple indicates those without sample overlap. PGSC - PGS Catalog; FG - FinnGen.

We first assessed the compatibility of genetic variants between the PGS models and the FinnGen dataset, which includes 21,342,234 variants (**Materials and Methods, PGS Catalog and FinnGen harmonization**). Of the 3,688 models available in the PGS Catalog, 3,025 (82%) had at least 75% variant overlap with FinnGen, and were selected for further analysis (**Fig. 1b**). The median number of variants in PGS models was 7,372 (min=1, max=10,318,272), and only ∼9% of models were based on HapMap3 variants that are often used for PGS model construction^14^ (**Fig. 1c**). Most of the analyzed models (73%) were published after 2022 (**Fig. 1d**).

We then harmonized phenotype definitions by matching PGS trait descriptions to FinnGen disease endpoints, defined using nationwide registries and the International Classification of Diseases (ICD) codes (https://risteys.finngen.fi/). While 1,717 models (out of 3,025) lacked a direct endpoint match often because they were derived for quantitative traits not available in FinnGen at the time of analysis, such as neuroimaging traits (e.g., brain volume, gray matter volume) or biomarker levels (e.g., blood proteins, creatinine, cystatin C) they were still retained for PGS calculations and further analyses (**Fig. 1e Supplementary Fig. 1c**).

We performed several rounds of manual review to flag and annotate models that had sample overlap with FinnGen Such overlap can lead to inflated performance estimates (**Supplementary Fig. 2**). We identified 460 models (15%) with present overlap (**Materials and Methods, PGS Catalog and FinnGen harmonization**). Each was flagged in the results database and omitted from all subsequent benchmarking and predictive model analyses. The remaining 2,565 models (85%), without sample overlap, were mapped to 227 unique FinnGen endpoints (**Supplementary Table 1**).

In total, we generated individual-level polygenic scores for 3,025 models from the PGS Catalog and 143 additional models developed by the FinnGen core team (see **Web resources, FinnGen core team PRS-CS pipeline**), all processed in the same way.

### Evaluation of PGS Catalog models using FinnGen cohort

We analyzed 779 PGS models matched to 221 FinnGen binary endpoints, omitting any with identified sample overlap. Each PGS was assigned to one of eight broad trait categories defined in the PGS Catalog (**Fig. 2a**)^8^ FinnGen endpoints were preprocessed to better reflect disease prevalence in the Finnish population (**Materials and Methods, FinnGen disease endpoints**). For each model, we quantified the ability to discriminate disease status with the area under the receiver-operating characteristic curve (ROC AUC), and assessed the significance of association with the corresponding endpoint using logistic regression, adjusting for sex, age, the first six principal components, and the genotyping array (**Fig. 2a**).

**Figure 2.**
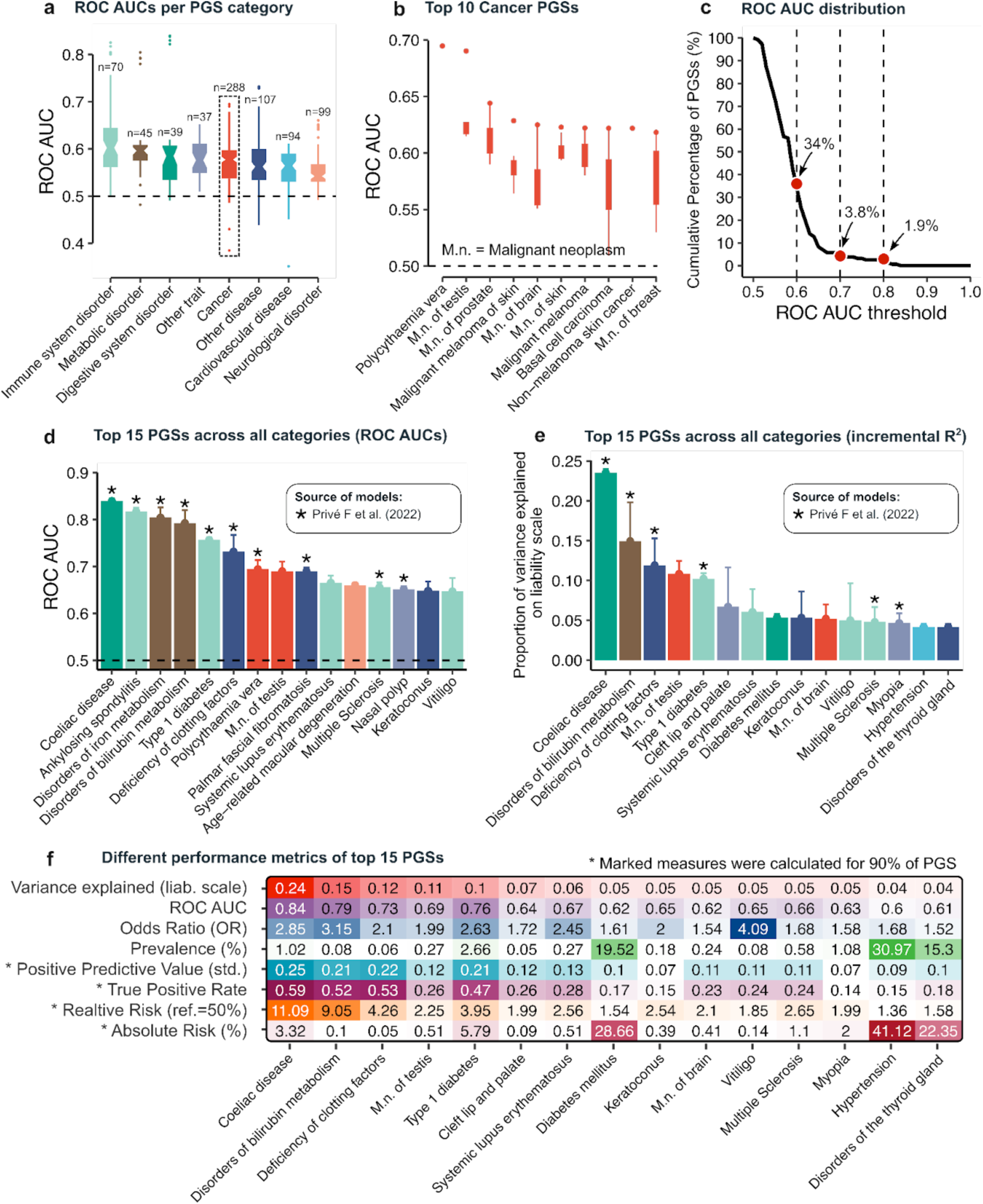
Evaluation of PGS Catalog models using the FinnGen cohort. (a) ROC AUC distributions for eight PGS categories based on PGS Catalog annotations. The dashed box highlights the ROC AUC distribution for cancer PGSs. (b) ROC AUC distributions for the top ten PGSs Multiple models available for each disease, with varying ROC AUCs The box plots show these distributions, and the red dot indicates the model achieving the highest ROC AUC for the selected disease. (c) The ROC AUC distribution for the 157 best-performing scores Red dots represent the percentage of scores exceeding the corresponding ROC AUC thresholds. (d) The top 15 PGSs across all categories ranked by unadjusted ROC AUC. Asterisks here and in (e) indicate PGSs derived from the publication by Privé F. et al. (PGP000263). (e) The top 15 PGSs across all categories ranked by the percentage of variability explained o the logistic liability scale. (f) Various performance metrics for these top 15 scores including standardized positive predictive value (set at 5% prevalence) and relative risk (using the 50th percentile as the reference). M.n., malignant neoplasm.

First, we sought to identify the most predictive PGSs for the corresponding clinical endpoints. For each endpoint, we evaluated all matched PGSs and selected the one that was significantly associated (p<1.06*10^-5^) with the trait and had the highest ROC AUC. Endpoints that lacked any significantly associated scores were excluded from the analysis. For example, among 13 PGSs for testicular cancer, PGS000796^15^ was significantly associated with the disorder (OR=1.77, CI:1.63-1.92) and achieved the highest ROC AUC of 0.69 (CI:0.67-0.71; **Fig. 2b**). Other top-performing PGSs in the Cancer category included polycythemia vera (PGS001810^16^ AUC=0.69; CI:0.68-0.71), prostate cancer (PGS002016^16^ AUC=0.64; CI:0.64-0.65), skin melanoma (PGS000766^17^ AUC=0.63; CI:0.62-0.64) and brain (PGS003737^18^ AUC=0.62; CI:0.61-0.64). Altogether, we identified 157 best-performing PGSs for the corresponding endpoints (**Supplementary Fig. 3; Supplementary Table 2**). Only six scores (3.8%) achieved a ROC AUC of 0.70 or higher (**Fig. 2c**). The highest-performing scores included those for coeliac disease (PGS001856^16^ AUC=0.84, CI:0.84-0.85), ankylosing spondylitis (PGS001876^16^ AUC=0.82, CI:0.81-0.83), disorders of iron metabolism (PGS001823^16^; AUC=0.80, CI:0.78-0.83), bilirubin metabolism (PGS002032^16^; AUC=0.79, CI:0.77-0.82), and type diabetes mellitus (PGS001817^16^ AUC=0.75, CI:0.75-0.76; **Fig. 2d**). Most of these top scores originated from Privé F et al. ^16^ were based on UK Biobank summary statistics, and generated with penalized regression.

We observed substantial individual-level variability among PGSs for the same disease, despite similar ROC AUCs. For example, two hypertension PGSs (PGS002701^19^ and PGS002047^16^) achieved equivalent ROC AUCs of 0.60 and 0.59, respectively (**Supplementary Fig. 4a,b**), yet shared only 30% overlap in identifying individuals in the top 2.5% of the PGS distribution (**Supplementary Fig. 4c-e**). This mismatch persisted even among top-performing scores For instance, four coeliac-disease models with ROC AUCs of 0.83-0.84 shared only 18% to 45% of individuals in the bottom 2.5% quantile, which ca be partly explained by the multimodal shape of the distributions (**Supplementary Fig. 5**). The main drivers of this individual-level discordance were the choice of GWAS summary statistics and the PGS-construction method when both scores were built from the same GWAS (**Supplementary Fig. 6**). These findings underscore the need to consider such individual-level variability when selecting or applying a single PGS for risk stratification^20,21^ To facilitate such comparisons, we provide a Spearman rank-correlation matrix for all 3,025 PGSs, capturing rank concordance between each pair of (**Data and code availability**).

For each best-performing PGS, we additionally calculated the variance explained on the logistic liability scale^22^ The top five scores by this metric were coeliac disease 23.5% (CI:23-24 %), disorders of bilirubin metabolism 15% (CI:12-20%), congenital vitamin K-dependent coagulation-factors deficiency 12% (CI:7-15 %), cancer of the testis 11% (CI:8-12%), and type diabetes mellitus 10% (CI:10-11%) (**Fig. 2e; Supplementary Fig. 7; Supplementary Table 4**).

Following the polygenic score reporting standards^7^ we computed additional metrics for each of the 779 PGSs, including the odds ratio (OR), true-positive rate (TPR), standardized positive predictive value (assuming 5% prevalence), and relative and absolute risks (**Fig. 2f; Supplementary Note, PGS evaluation measures; Supplementary Table 5**). For example, coeliac disease demonstrated the highest TPR of 0.59 (at the 90th-percentile threshold), a relative risk of 11.09 (90th vs 50th percentile), and an OR of 2.85, with an absolute risk of 3.32%, consistent with its 1% prevalence in FinnGen.

Further, we evaluated each PGS separately within each major ancestry group in FinnGen. Of the 473,681 FinnGen participants, 19,947 were non-Finnish: 15,101 non-Finnish Europeans, 1,620 Admixed Americans, 690 East Asians, 433 Africans, 352 South Asians, and 1,751 individuals classified as “Others” (**Supplementary Table 3; Materials and Methods, Ancestry Prediction**). Endpoints with fewer than eight cases in a given ancestry were excluded, reducing the analysis to 740 scores and 200 corresponding endpoints. Because case counts differed ancestries, the number of endpoints tested for each group varied accordingly (**Supplementary Fig. 8**). In total, we identified 122 best-performing scores for non-Finnish Europeans, 54 for Admixed Americans, 16 for East Asians, 9 for Africans, 14 for South Asians, and 55 for “Others.” Across ancestry groups, immune-related and metabolic traits consistently achieved the highest predictive performance (**Supplementary Fig. 9**). Notably, PGSs for type 2 diabetes PGS002720^19^ (ROC AUC range across ancestries: 0.57-0.64) and PGS000807^23^ (ROC AUC range: 0.59-0.64) were the only scores to achieve statistically significant discrimination in every ancestry group examined.

### Atlas of PGS-based Phenome-Wide Association Studies

We performed a PheWAS for 3,168 polygenic scores across 4,739 FinnGen endpoints, employing four complementary designs (**Fig. 3a; Materials and Methods, Phenome-Wide Association Study**): (1) “intact” (n=3,168); (2) “survival” (n=3,168), in which we utilized the Cox proportional-hazards model instead of logistic regression; (3) “exclusion” (n=1,172), in which we excluded individuals with the target phenotype from the analysis to mitigate phenotypic hitchhiking ^5^; and (4) “noMHC” (n=3,023), in which we removed all variants located in the MHC locus from each PGS model. The “exclusion” and “noMHC” designs provided important validation and interpretative support for the primary “intact” analysis^24^

**Figure 3.**
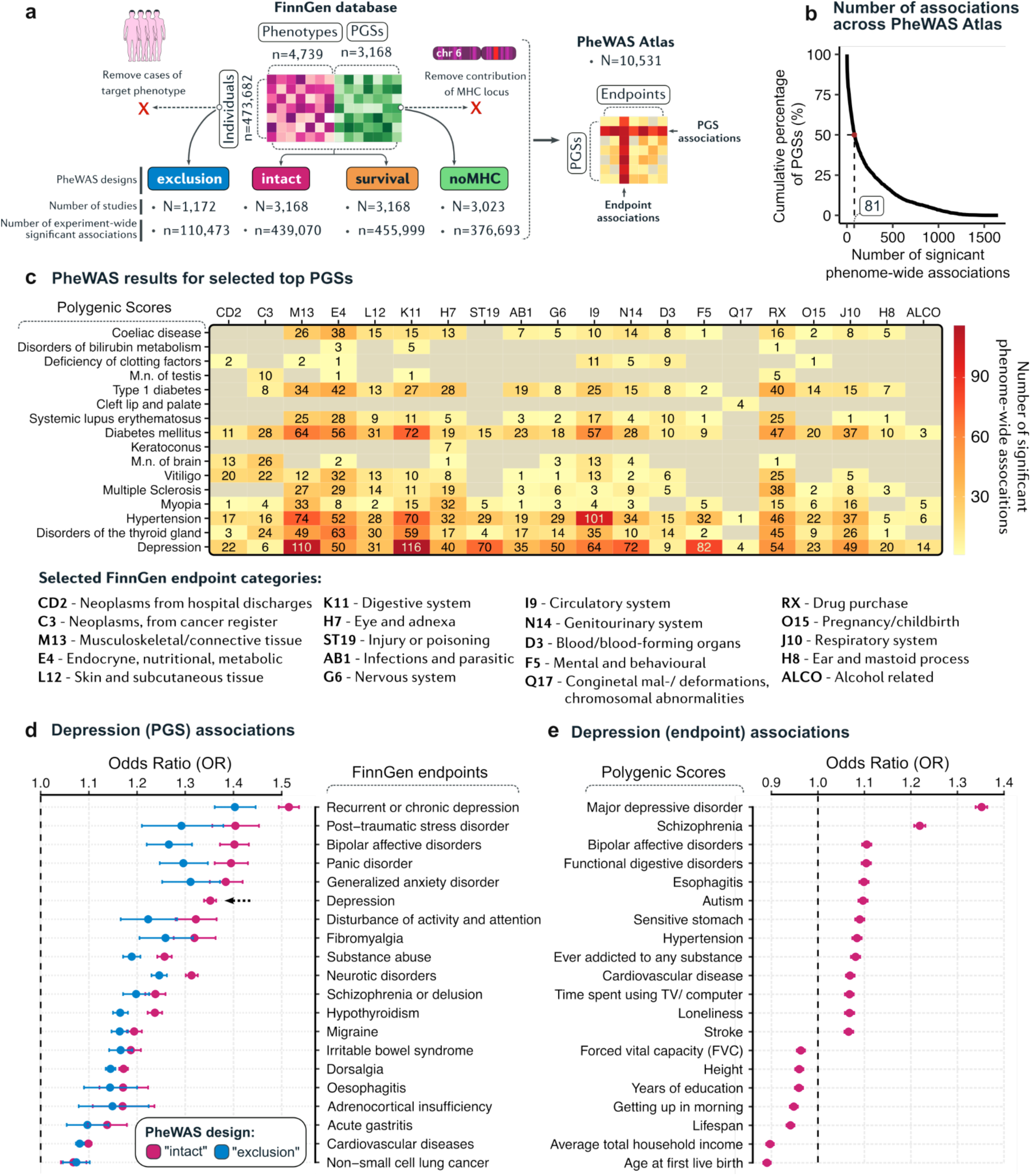
Atlas of PGS-based phenome-wide association studies. (a) Overview of the four distinct PheWAS designs used, along with the corresponding numbers of studies and experiment-wide significant associations In the “exclusion” design, cases of the target phenotype were excluded to mitigate confounding effects on secondary associations. In the “noMHC” design, variants in the MHC region were removed from the PGS model to evaluate PGS–endpoint associations based solely on non-MHC variants. (b) Number of significant phenome-wide associations across the PheWAS Atlas; the red dot indicates the median number of associations. (c) PheWAS results for the top 15 PGSs and Depression. Numbers and color gradients represent the number of significant phenome-wide associations per FinnGen endpoint category. (d) PheWAS results for the best-performing PGS for Depression (PGS000907). Crimson indicates results from the “intact” PheWAS design, while blue represents results from the “exclusion” design. The black asterisk highlights the PheWAS result for the target Depression endpoint (F5_DEPRESSIO). The blue bar is absent because F5_DEPRESSIO cases were removed in the “exclusion” design. (e) Combined PheWAS results for the Depression endpoint, showing associated non-target PGSs.

In the “intact” design, 3,001 out of 3,168 PGSs had at least one phenome-wide significant association (Bonferroni threshold was determined for every PGS as p 0.05/4,739, **Materials and Methods, Phenome-Wide Association Study**). The median number of associations per score was 8 (mean=206, max=1,632; **Fig. 3b**). Endpoints were grouped using ICD-10-based tags curated by the FinnGen team (e.g., I9 circulatory-system disease; **Supplementary Table 6**). Most scores showed the strongest associations within their target trait category, but often revealed additional associations in other domains (**Fig. 3c**). Through this analysis, we have created the PGS-PheWAS atlas, which contains information on each PGS-endpoint association in FinnGen.

The PGS-PheWAS atlas can be used to investigate clinical endpoints associated with a specific PGS. For example, the top-performing PGS for depression (PGS000907^25^) was strongly associated with the FinnGen depression endpoint (F5_DEPRESSIO; OR=1.35, CI:1.34-1.36) and showed an additional 1,256 significant associations (**Fig. 3c-d**). These included endpoints related to digestive-system disorders (n=116), musculoskeletal and connective-tissue disorders (n=110), and mental-health conditions (n=82). The most associated traits included bipolar disorder (OR=1.40, CI:1.37-1.43), schizophrenia or delusion (OR=1.31, CI:1.29-1.34), panic disorder (OR=1.39, CI:1.36-1.43), and attention-deficit/hyperactivity disorder (OR=1.32, CI:1.28-1.36). Notably, the depression PGS was also significantly associated with hypothyroidism (OR=1.24, CI:1.22-1.25), irritable bowel syndrome (OR=1.19, CI:1.17-1.21), and cardiovascular disease (OR=1.10, CI:1.09-1.11), consistent with earlier studies linking depression to each of these conditions ^26–30^

To assess whether secondary associations were driven by the primary trait itself or MHC region, we compared “exclusion” and “noMHC” with the “intact” results^24^ n this case, MHC-locus removal had no effect on phenotypic associations as the original PGS did not include MHC variants. Removal of depression cases in the “exclusion” design reduced effect sizes but previously described associations remained significant, confirming their robustness (**Fig. 3d**).

Alternatively, the PGS-PheWAS atlas be used to identify which PGSs, beyond the for depression, are significantly associated with the corresponding FinnGen endpoint. For example, the PGS for schizophrenia (OR=1.22, CI:1.20-1.23), bipolar affective disorder (OR=1.10, CI:1.09-1.12), esophagitis (OR=1.10, CI:1.09-1.11), and hypertension (OR=1.08, CI:1.07-1.09) were significantly associated with the depression endpoint (F5_DEPRESSIO). In contrast, significant negative associations were observed for polygenic scores related to the age at first live birth (OR=0.89, CI:0.88-0.90), average total household income (OR=0.89, CI:0.89-0.91), lifespan (OR=0.94, CI:0.93-0.95), years of education (OR=0.95, CI:0.95-0.97), and height (OR=0.95, CI:0.95-0.97; **Fig. 3e**).

We provide to the entire atlas of 10,531 PheWASs, including all study designs, via web application (see **Data and code availability**).

### Integration of multiple PGSs improves disease prediction

To identify traits that could be predicted more efficiently using a combination of PGSs, it was critical to ensure that there was no undetected sample overlap between the score-development cohorts and FinnGen ^3^ Therefore, we limited our analysis to PGS models from six publications, covering nearly half of the PGS Catalog, and manually verified that none utilized FinnGen samples (**Fig. 4a**). From these studies, we extracted 1,514 PGS models covering 110 diseases that matched FinnGen endpoints. For each disease, we then tested whether combining multiple scores could improve predictive accuracy.

**Figure 4.**
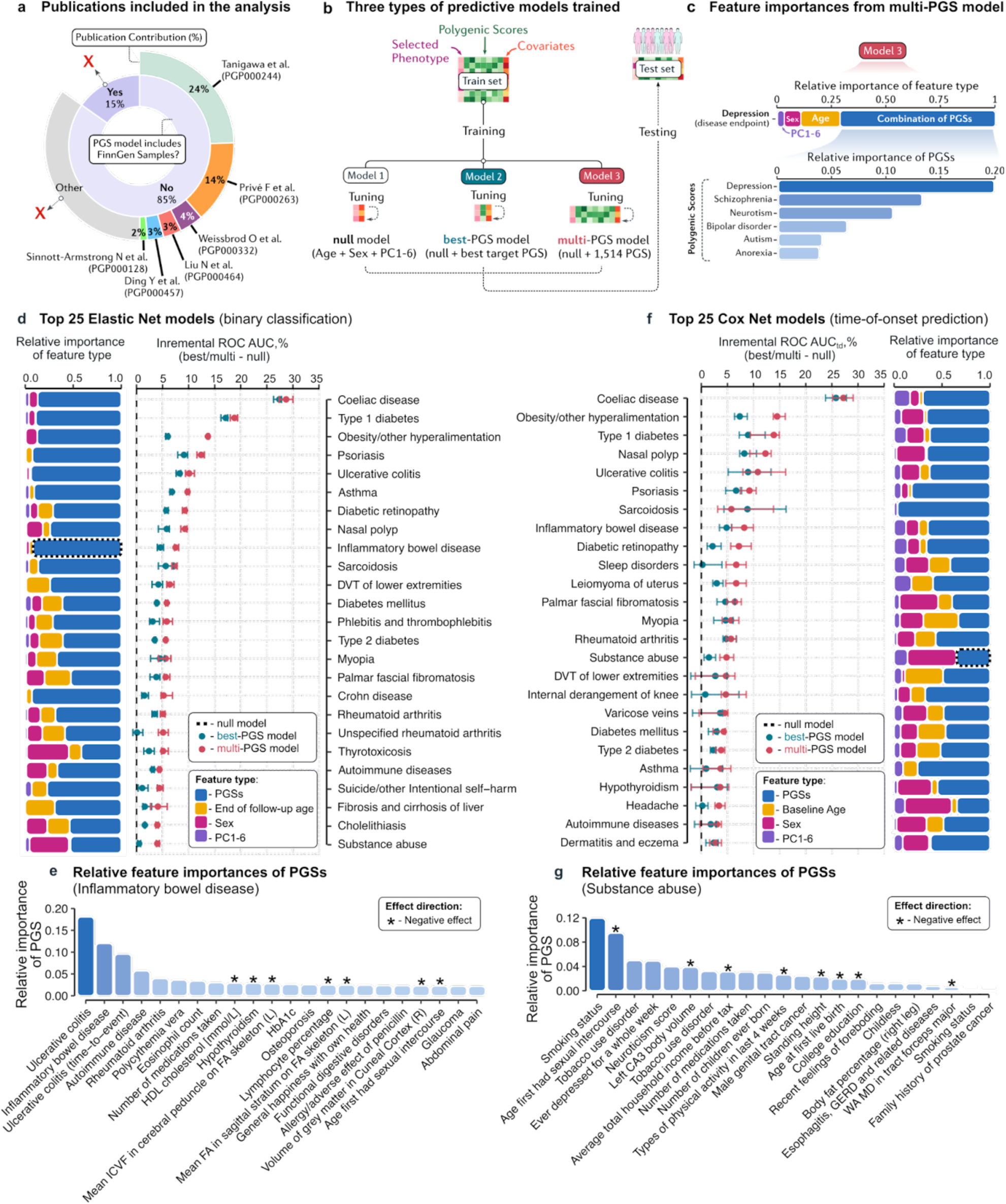
Integration of multiple PGSs improves disease prediction. (a) Six publications were kept for further analysis, covering approximately 50 % of the retained PGS Catalog models and reliably lacking FinnGen sample overlap. (b) A scheme of the training and testing process for the three predictive model types used in subsequent experiments. (c) Representation of the relative feature importances visualized in the following panels. (d) Top 25 Elastic Net models for binary disease-status classification. The left panel shows incremental ROC AUC improvements (percentile scale) between best/multi and null models. The right panel displays relative feature importances for four feature types: blue (all PGSs combined), yellow (age at the end of follow-up), crimson (sex), and violet (first six principal components). The black dashed box highlights PGS feature importances for inflammatory bowel disease (IBD). (e) Relative feature importances for the optimal combination of PGSs for IBD. (f) Top 25 Cox Net models for time-to-event prediction. The left panel shows incremental time-dependent ROC AUC (1-10 years) improvements between best/multi and nul models In this case yellow represents baseline age. The black dashed box highlights PGS feature importances for substance abuse (g) Relative feature importances for the optimal combination of PGSs for substance abuse Asterisks in (e) and (g) indicate scores with a negative effect on the disease endpoint.

FinnGen was split into training and testing sets (**Fig. 4b**). In the training set, we built three models: a null model (sex, age, and six genetic PCs), a best-PGS model (null the top single PGS), and a multi-PGS model (null a optimal combination of PGSs). Scores with on-zero coefficients, given by the elastic net, were considered informative (**Fig. 4c**). Elastic-net logistic regression was used for binary outcomes, and elastic-net Cox proportional-hazards models for time-to-event prediction (**Materials and Methods, Risk prediction models**).

For disease-status prediction, 80 of 110 endpoints (73%) showed a significant ROC AUC improvement when the best single PGS was added to the null model (mean ΔAUC=0.027, p<2.2*10^-16^ **Fig. 4d–f; Supplementary Fig. 10; Supplementary Table 7**). The most substantia gains were observed for coeliac disease (ΔAUC = 0.27) and type diabetes (ΔAUC = 0.17; **Fig. 4d**). The multi-PGS models showed improved performance for 10 of 110 models (mean ΔAUC increase 0.037, p<2.2*10^-16^), with coeliac disease and type diabetes achieving an improvement of 0.29 and 0.19, respectively. Comparing the best-PGS to multi-PGS models revealed performance gains for 87 endpoints (mean ΔAUC = 0.017, p<2.2*10^-16^), including obesity and other hyperalimentation (ΔAUC = 0.08), rheumatoid arthritis (ΔAUC = 0.05), and Crohn’s disease (ΔAUC=0.04; **Supplementary Fig. 11**). These improvements largely driven by a combination of several target and non-target PGSs. In the inflammatory bowel disease (IBD) model, for example, the most influential target scores came from IBD itself and ulcerative colitis, while non-target scores came from rheumatoid arthritis, polycythemia vera and eosinophil count (**Fig. 4e**).

We applied the same approach to predict time-to-event by fitting elastic-net Cox models (**Supplementary Table 8**). Compared with a null model, the best PGS delivered a significant increase in time-dependent ROC AUC for 29 of 110 endpoints (26%; mean ΔAUC=0.048, p<2.2*10^-16^). Incorporating multiple PGSs raised this to 42 endpoints (38%; mean ΔAUC=0.056, p<2.2*10^-16^), with the largest gains for coeliac disease (ΔAUC=0.29), obesity and related hyperalimentation disorders (ΔAUC=0.15), and type diabetes (ΔAUC=0.11) (**Fig. 4f; Supplementary Fig. 12**). When we directly compared multi-PGS models with their best-PGS analogues, 15 endpoints (13%; mean ΔAUC=0.037, p<2.2*10^-16^) showed further improvement. The biggest improvements were observed for obesity (ΔAUC=0.08), sleep disorders (ΔAUC=0.05), and diabetic retinopathy (ΔAUC=0.04; **Supplementary Fig. 13**). These gains were also mainly driven by non-target PGSs. As an example, substance-abuse onset prediction benefited most from the integration of scores for smoking status, age at first sexual intercourse, week-long depressive episodes, and a neuroticism score (**Fig. 4g**).

For all 110 diseases, we provide a list of contributing PGSs and their corresponding weights from the multi-PGS model for both disease status and time-to-event prediction (see **Supplementary Tables 9,10**).

### Reference PGS distributions and predictive models

To enable external use while preserving data-privacy regulations, we parameterized each PGS distribution into shareable percentiles. These percentiles allow the distribution to be reconstructed and used reference for risk stratification. To prevent bias in percentile interpretation due to population structure, we ancestry-adjusted each of the 3,168 scores, following established methodology^32^

Each residualized genetic principal components (PCs) to population-structure effects, making the distributions comparable across different ancestries (**Fig. 5a-b**). For each PGS, we first computed predicted scores based on genetic PCs alone. Some PGSs, such as those for height (r=0.62; PGS002989^6^), intracrania aneurysm (r=0.59; PGS003407^33^), and atopic dermatitis (r=0.54; PGS002755^34^), showed a substantial correlation with population structure (**Fig. 5c Supplementary Table 11**). We then computed adjusted PGSs by subtracting the *predicted* component from the *raw* score This effectively eliminated ancestry-driven variation, as the adjusted scores were uncorrelated with PCs (**Supplementary Fig. 14**). For example, the raw breast-cancer PGS (PGS000001^35^) overestimated percentile assignments in individuals of African ancestry (**Fig. 5e**), but the adjusted PGS resulted in comparable percentile assignments across ancestries (**Fig. 5f**).

**Figure 5.**
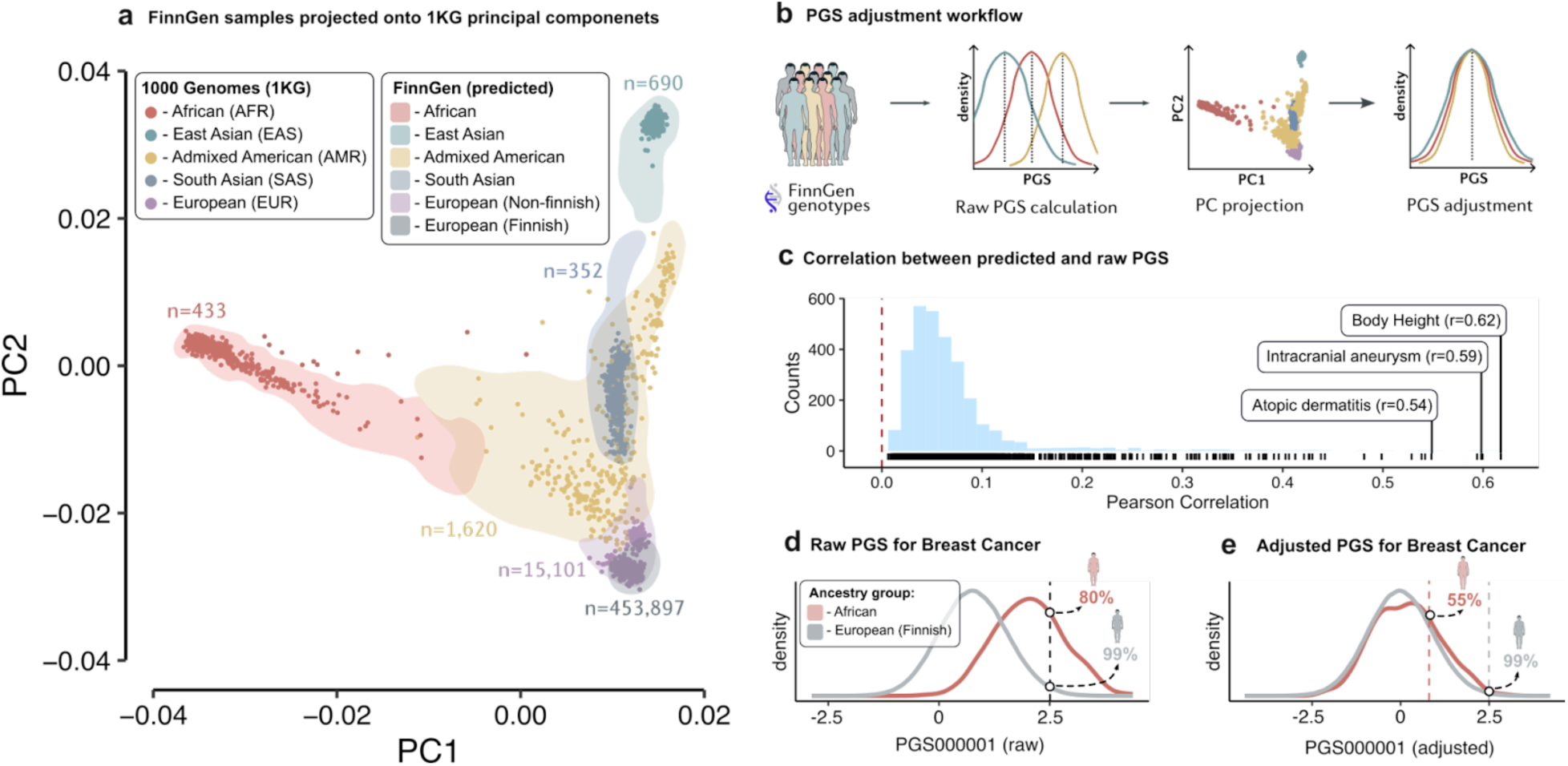
Ancestry-adjusted polygenic scores. (a) Projection of 473,681 FinnGen samples onto the first two principal components derived from the 1000 Genomes Project. Colored points represent 1000G reference samples, and shaded areas indicate the distribution of FinnGen samples by inferred ancestry. (b) Overview of the PGS-ancestry-adjustment procedure. (c) Correlation between raw PGS values and predicted PGS values, representing the portion of genetic risk explained solely by population structure; the dashed brown line represents zero correlation. (d) Breast-cancer PGS distributions for Finnish and African ancestry groups highlighting how a given aw PGS value corresponds to different percentiles across populations due to population structure. (e) Adjusted breast-cancer PGS distributions for the populations. After adjustment, the distributions are better aligned, and individual scores become more interpretable across ancestries.

Within each ancestry group we recalculated ROC AUCs for the ancestry-adjusted scores and compared them with ROC AUCs for the raw PGSs (**Supplementary Table 3**). Only a few scores exhibited a statistically significant difference in ROC AUC, with the highest number observed in the African ancestry group (n=37), followed by Admixed Americans (n=20) and East Asians (n=15) (**Supplementary Fig. 15**).

To further enhance the utility of the provided resources, we incorporated ancestry-adjusted PGSs into time-to-event models. Because the adjustment removes the need for explicit principal-component covariates, these models can be applied externally with minima inputs only the individual’s sex, current age, and PGS percentile, which can be calculated locally (**Supplementary Note, External predictive models**). Twenty-two best-PGS models met our performance criteria each achieved a time-dependent ROC AUC above 0.65, displayed excellent calibration (D-calibration^36^ p>0.99), and improved ΔAUC by at least 0.01 over a null model containing only sex, age, and PCs (**Supplementary Fig. 16; Materials and Methods, Risk Prediction Models**).

To facilitate interpretation of PGS values from external cohorts, we developed a framework that visualizes user-provided scores against the ancestry-adjusted reference distributions, and enables high-throughput, customizable selection of individuals with specific genetic profiles (**Data and Code Availability**).

### Polygenic Score browser (PGS browser)

All PGS models matched to the FinnGen cohort (n=3,168), along with results from phenome-wide association studies (n=10,540), PGS Catalog model evaluations, ancestry-adjusted reference distributions (n=3,168), and time-to-event predictive models (n=22) were made accessible through a dedicated web application the PGS Browser. (**Fig. 6; Data and Code Availability**).

**Figure 6.**
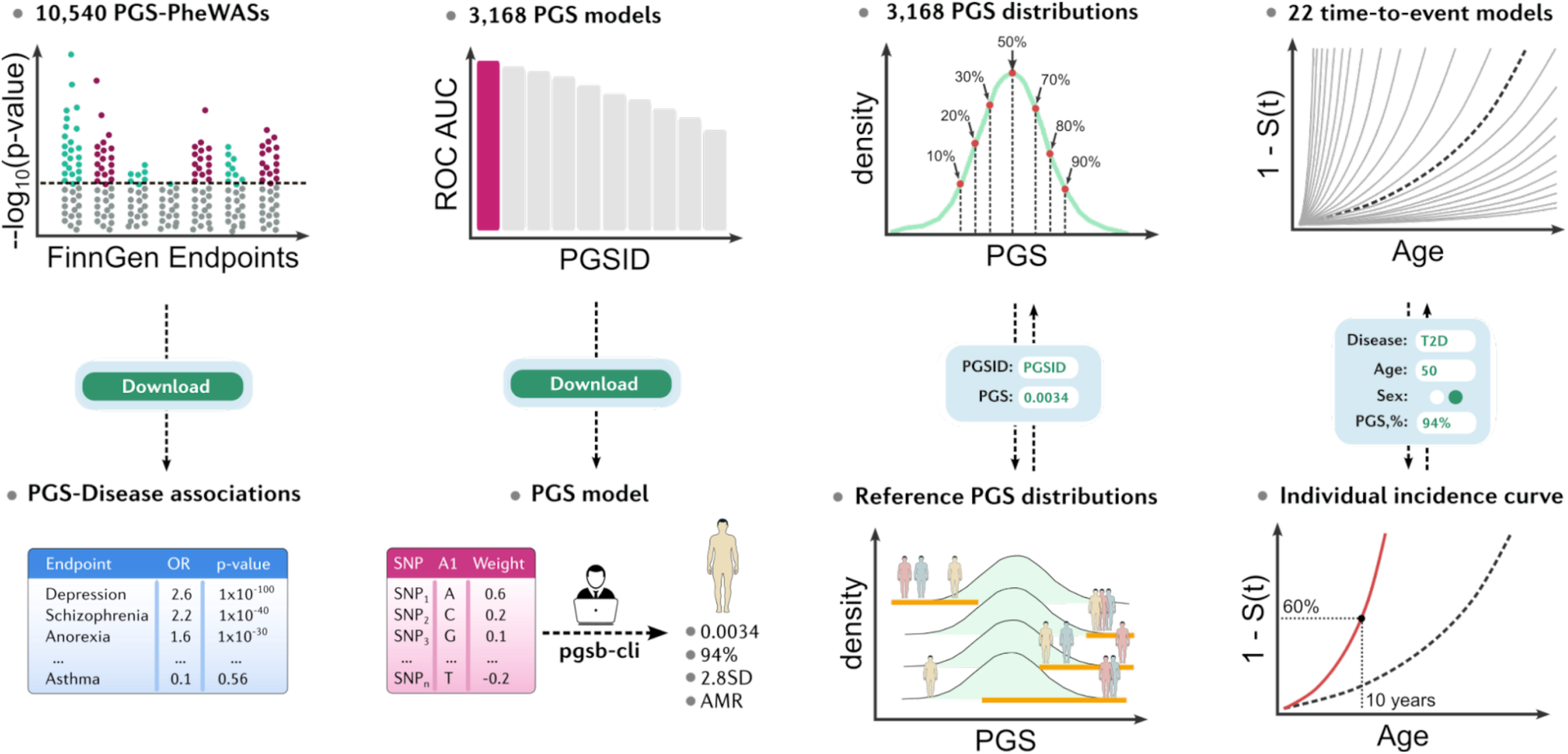
Polygenic Score Browser (PGS Browser). An overview of the data types and functionalities available in the PGS Browser the top leve llustrates the available data and predictive models, the middle level highlights elements of the graphical interface, and the bottom level shows the results displayed to the user. Pgsb-cli - pgs browser command-line interface.

We also provide “pgsb-cli” Docker-based command-line tool that locally. Given PGS model downloaded from the PGS Browser and a patient’s genotype data, it automatically matches variants, computes the ancestry-adjusted PGS, and reports standardized values and percentiles. Only these aggregated percentiles need to be sent to the web interface for predictive models, keeping all raw genetic data on the user’s machine (**Fig. 6**).

Importantly, of these subject to data-sharing restrictions. The open provided by the PGS Browser enables researchers outside FinnGen to explore, analyze, and apply these polygenic score resources and tools, ensuring equitable access and amplifying their translational impact.

## Discussion

In this study, we benchmarked the performance of 3,168 polygenic score models using a dataset of 473,681 FinnGen participants. Such a systematic evaluation of most of the currently publicly available PGS models within a single, biobank-scale cohort has enabled reliable cross-model comparisons and highlighted both technical and practical challenges. Over the past decade, the UK Biobank has supplied the GWAS data for nearly 72% of all publicly released PGS models, making independent performance evaluation difficult. FinnGen offers compelling alternative: it matches the UK Biobank’s breadth of phenotypes, yet provides larger case numbers for the majority of disorders and shares little sample overlap with the GWAS cohorts used to construct PGS models. Thus, FinnGen is uniquely suited to deliver unbiased estimates of these models’ predictive accuracy and translational value.

Large-scale PGS analyses demand extensive automation, which, n turn, depends standardized data formats and reliable metadata. The PGS Catalog largely meets these needs: its submission standards and detailed annotations make most scores readily accessible and reusable Nonetheless, we encountered integration challenges arising from occasional discrepancies in reference alleles, effect sizes, and trait descriptions. The most consequential obstacle was hidden sample overlap with FinnGen, which inflated performance metrics. In practice, any PGS showing a target-trait AUC 0.65 often traced back to unreported nclusion of FinnGen (or its legacy cohorts) in the source GWAS, which had to be verified manually in the original publications. Thus, even with current Catalog guidelines, cohort reporting, particularly for scores derived from GWAS meta-analyses, can remain incomplete. However, this issue is a greater concern for cohort-based PGS studies than for translational or clinical applications, where participant recruitment is typically independent of GWAS research efforts.

Evaluation of the PGS Catalog revealed that only a small subset of models achieved a ROC AUC above 0.7. The majority of top-performing models were immune-related, were derived from Privé et al.^16^ and relied on UK Biobank summary statistics, reflecting both the extensive phenotype coverage and the cohort’s predominantly European origin. We provide detailed performance statistics for each PGS model, which be used to their translational potential and guide genetic testing.

From a translational perspective, we found considerable variability in individual risk estimates among PGSs for the same trait. This heterogeneity poses a reproducibility challenge but also creates an opportunity for improving prediction quality. Variability arises from methodological differences (e.g., GWAS source, PGS algorithm, development set, parameters chosen) and from the stochastic nature of Markov chain Monte Carlo–based methods (e.g. PRS-CS^37^ LDPRed2^38^ etc.). To improve reproducibility, we suggest using a single, consistent model rather than relying on any score with similar global metrics. At the same time, informative scores that are only moderately correlated be combined to boost accuracy, shown for inflammatory bowel disease. We provide a correlation matrix and ROC AUC values for all binary-trait scores giving a reference for the expected level of individual-level discordance within each trait.

The potential of multi-PGS modeling extends beyond scores for the target trait itself. Our phenome-wide association analyses revealed umerous associations between disease endpoints and PGSs for genetically related traits, suggesting that shared architecture can be harnessed to improve prediction performance. Among the 110 diseases, multi-PGS models consistently outperformed single-score models, yielding notable AUC gains for obesity, rheumatoid arthritis, Crohn’s disease, and several other conditions. As large-scale PGS pipelines^39^ and score-integration frameworks^40,41^ continue to improve, such cross-trait, multi-PGS approaches should become even more practical and efficient.

To facilitate the integration of all presented findings into the analysis of external cohorts, we generated ancestry-adjusted reference distributions for all 3,168 PGSs in FinnGen and made them publicly available through the PGS Browser. Researchers can visualize each distribution, contextualize individual scores, browse PheWAS results, apply time-to-event prediction models, and construct custom cohorts by specifying flexible multi-PGS nclusion or exclusion criteria Nevertheless, there is a practical caveat, even minor differences in the variants used to calculate a score for a new dataset can shift its distribution relative to the FinnGen reference. We therefore recommend a stringent variant-matching threshold (≥95%); when that is unattainable, users should either substitute linkage-disequilibrium proxies for missing variants or choose an alternative PGS.

Extending polygenic risk assessment to resource-limited and underserved settings demands FAIR, biobank-scale resources delivered through user-friendly, secure platforms that lower technical barriers and ensure equitable benefit sharing^42–44^ The PGS Browser meets these requirements by offering publicly accessible, biobank-linked PGS data and tools, expanding the translational potential of polygenic scores and engaging the global research and clinical community.

## Materials and Methods

### Study cohort

The dataset includes 473,682 Finnish individuals from FinnGen Release 11, containing both genetic and clinical data. Details about the cohort and study protocols have been previously documented ^45^ Experimental findings, predictive models, and PGS distributions are shared under the authorization of the FinnGen Scientific Committee (project number F_2020_073). All participants provided written informed consent.

### Genotyping and imputation

The FinnGen samples were genotyped using Illumina and Affymetrix arrays (Illumina and Thermo Fisher Scientific). These samples underwent standardized quality control, imputation, and post-imputation processing steps outlined in the previously described protocol (see **Web resources**).

### PGS Catalog and FinnGen harmonization

Variants from each of the 3,688 PGS models were downloaded, matched to FinnGen variant data, and aggregated into effect-size matrices using PGS Catalog utilities (see **Supplementary Note, PGS Models Processing**). We evaluated all models in their original form, intentionally avoiding the substitution of unmatched variants with proxies in strong linkage disequilibrium. This decision helped prevent the introduction of additional variability and allowed for a direct assessment of each model’s original performance.

PGS models developed from cohorts that potentially overlapped with FinnGen or its legacy cohorts (as reported in the GWAS) flagged accordingly (see **Supplementary Table 12**).

Individual PGS scores were calculated using PLINK 2.0’s *score* function ^46^ Reported traits for each model were matched to FinnGen endpoint descriptions via regular expressions, and all matches were manually confirmed.

### Ancestry Prediction

FinnGen includes 453,734 participants of Finnish ancestry and 19,947 participants of non-Finnish ancestry, often treated as PCA outliers and excluded from prior analyses. To characterize them, we projected al ndividuals into the 1000 Genomes PCA space and used a gradient-boosting classifier trained the first three PCs (10-fold cross-validation) to assign continental ancestry. Samples with more than 65% posterior probability were labelled accordingly; all others were marked “Others” (see **Supplementary Note, Ancestry prediction in FinnGen cohort**).

### FinnGen disease endpoints

From FinnGen Release 11, we utilized 4,739 endpoints, each with more than six cases These endpoints represent binary features indicating the presence or absence of specific conditions, with diagnoses made on the basis of the International Classification of Diseases (ICD-8, ICD-9, and ICD-10) criteria. Detailed inclusion/exclusion criteria and case/control definitions are available via the Risteys portal (see **Web Resources**). All endpoints were grouped into 38 categories, with corresponding tags (e.g., I9, F5), as defined by the FinnGen core team (**Supplementary Table 6**). For PGS evaluation and predictive modeling, samples excluded from the control set were reintroduced to better reflect the control population and disease prevalence in the Finnish population. For PheWAS experiments, we kept the endpoints intact.

### Phenome-Wide Association Study

For each PheWAS, we fitted a logistic-regression model with LDLT decomposition using the *fastglm* R package to speed up computations ^47^ We adjusted the relationship between the standardized PGS and each endpoint for sex age at the end of follow-up, PCs 1-6, and six genotyping-array dummy variables:

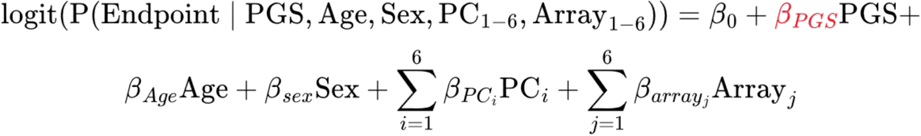

Our primary focus was β*PGS*, with other factors included to address potential residual confounding. Additionally, we computed unadjusted ROC AUC for each PGS-endpoint pair using the *bigsnpr* R package ^48^ For gender-specific traits, we excluded β*_sex_Sex* To handle multiple comparisons for assessment of associations for a single PGS, we set a Bonferroni threshold at p-value<1.06*10^-5^ (0.05/4,739). The same type of analysis was described previously ^49^ When determining the experiment-wide number of significant associations we have used Bonferroni adjustment for the total number of comparisons 0.05/(4,739 Endpoints 3,168 PGS models). In addition, we performed three alternative PheWAS designs: (1) “exclusion” in which individuals with the target phenotype were removed to mitigate confounding in secondary associations ^5^ (2) “noMHC” in which variants in the MHC region (chr6: 28.5-33.5 Mb, GRCh38) excluded and recalculated before conducting PheWAS the altered scores Comparing these results with the original design (unaltered scores and binary endpoints) allows s to identify associations driven solely by the MHC locus or phenotypic hitchhiking; and (3) “survival” in which logistic regression replaced with Cox proportional-hazards models (*survival* R package) to use time-to-event as the response variable.

Associations involving PGSs with sample overlap with Finnish cohorts were excluded from the main results but are provided, with full annotation, in the public data release (see **Data and code availability**).

### Risk prediction models

For binary classification, we used logistic regression with a elastic-net penalty, implemented in the *scikit-learn* Python package (v1.5.1) (see **Web Resources**). Regularization strength (C) and the elastic-net mixing parameter (l1 ratio) were optimized through grid-search cross-validation (cv=5), using ROC AUC as the scoring metric.

For time-to-event prediction, we utilized Cox’s proportional-hazards model with an elastic-net penalty, implemented in the *scikit-survival* Python package (v0.23.2) (see **Web Resources**). Follow-up years were used as the time scale, calculated by subtracting baseline age from either event age the end of follow-up. For each trait, individuals with baseline ages greater than event ages were excluded. Regularization strength (alpha) and the elastic-net mixing parameter (l1 ratio) were optimized using grid-search cross-validation (cv=5), with mean time-dependent ROC AUC (1-10 years) as the scoring metric.

Feature importances were derived from the weights of the elastic-net models. As all features were standardized before model fitting, their importances were directly comparable (**Supplementary Note, Disease Risk Prediction Models**).

To compare performance differences between sets of models (e.g., “null” vs “multi”), we used a paired Wilcoxon signed-rank test from the R *stats* package (v4.4.2).

### Graphical interface

The web application was primarily created using the *Shiny* R package (v1.10.0) (see **Web resources**). Unless specified otherwise, all analyses and software development were conducted using R 4.3.2.

## Ethics Statement

Individuals in FinnGen provided informed consent for biobank research, based o the Finnish Biobank Act Alternatively, separate research cohorts, collected prior the Finnish Biobank Act came into effect (in September 2013) and start of FinnGen (August 2017), were collected based on study-specific consents and later transferred to the Finnish biobanks after approval by Fimea (Finnish Medicines Agency) and the National Supervisory Authority for Welfare and Health. Recruitment protocols followed the biobank protocols approved by Fimea. The Coordinating Ethics Committee of the Hospital District of Helsinki and Uusimaa (HUS) statement number for the FinnGen study is Nr HUS/990/2017. Further details on permit and biobank decision numbers are available in the FinnGen Ethics Statement in the supplemental information.

## Data and code availability

All of the described results, PGS models, PheWASs, and predictive models and tutorials are available within PGS Browser: https://www.pgs.nchigm.org

Pgsb-cli: https://github.com/ArtomovLab/PGS_Browser

## Web resources

FinnGen consortium: https://www.finngen.fi/en/access_results

FinnGen imputation protocol: https://doi.org/10.17504/protocols.io.nmndc5e

Risteys R11 portal: https://r11.risteys.finregistry.fi/

PGS catalog: https://www.ebi.ac.uk/gwas/home

PGS catalog utilities: https://github.com/PGScatalog/pgscatalog_utils

FinnGen core team PRS-CS pipeline: https://github.com/FINNGEN/CS-PRS-pipeline

Shiny R package: https://github.com/rstudio/shiny

Scikit-learn Python package: https://scikit-learn.org/1.5/index.html

Scikit-survival Python package: https://scikit-survival.readthedocs.io/en/latest/

## Acknowledgments

This project was supported by funding from the Aging Biology Foundation to Artomov Lab.

We want to acknowledge the participants and investigators of FinnGen study. The FinnGen project is funded by two grants from Business Finland (HUS 4685/31/2016 and UH 4386/31/2016) and the following industry partners: AbbVie Inc., AstraZeneca UK Ltd, Biogen MA Inc., Bristol Myers Squibb (and Celgene Corporation & Celgene International II Sàrl), Genentech Inc., Merck Sharp & Dohme LCC, Pfizer Inc., GlaxoSmithKline Intellectual Property Development Ltd., Sanofi US Services Inc., Maze Therapeutics Inc., Janssen Biotech Inc, Novartis Pharma AG, and Boehringer Ingelheim International GmbH. Following biobanks are acknowledged for delivering biobank samples to FinnGen: Auria Biobank (www.auria.fi/biopankki), THL Biobank (www.thl.fi/biobank), Helsinki Biobank (www.helsinginbiopankki.fi), Biobank Borealis of Northern Finland (https://www.ppshp.fi/Tutkimus-ja-opetus/Biopankki/Pages/Biobank-Borealis-briefly-in-English.as px), Finnish Clinical Biobank Tampere (www.tays.fi/en-US/Research_and_development/Finnish_Clinical_Biobank_Tampere), Biobank of Eastern Finland (www.ita-suomenbiopankki.fi/en), Central Finland Biobank (www.ksshp.fi/fi-FI/Potilaalle/Biopankki), Finnish Red Cross Blood Service Biobank (www.veripalvelu.fi/verenluovutus/biopankkitoiminta), Terveystalo Biobank (www.terveystalo.com/fi/Yritystietoa/Terveystalo-Biopankki/Biopankki/) and Arctic Biobank (https://www.oulu.fi/en/university/faculties-and-units/faculty-medicine/northern-finland-birth-cohor ts-and-arctic-biobank). All Finnish Biobanks are members of BBMRI.fi infrastructure (www.bbmri.fi). Finnish Biobank Cooperative - FINBB (https://finbb.fi/) is the coordinator of BBMRI-ERIC operations in Finland. The Finnish biobank data can be accessed through the Fingenious® services (https://site.fingenious.fi/en/) managed by FINBB.

## Conflicts of interest

A provisional patent has been filed with respect to the findings described in this manuscript (USPTO serial no. 63/716,306).

Mark J. Daly is a founder of Maze Therapeutics.

## Author Contributions

Study design and conceptualization – NK, MPR, MJD, MA

Data analysis – NK, MPR, PDBP, MIK, VL, TPS, IM, MA

Statistical model design – NK, MA

Web application design – NK

Web application hosting support – AH

Manuscript writing – NK, MA

Funding acquisition – MA, MJD, AP

Project supervision – MA

Manuscript editing and approval – all authors

## Notes

The conflict of interest statement is at the end of the manuscript

### Author Declarations

Individuals in FinnGen provided informed consent for biobank research, based on the Finnish Biobank Act. Alternatively, separate research cohorts, collected prior the Finnish Biobank Act came into effect (in September 2013) and start of FinnGen (August 2017), were collected based on study-specific consents and later transferred to the Finnish biobanks after approval by Fimea (Finnish Medicines Agency) and the National Supervisory Authority for Welfare and Health. Recruitment protocols followed the biobank protocols approved by Fimea. The Coordinating Ethics Committee of the Hospital District of Helsinki and Uusimaa (HUS) statement number for the FinnGen study is Nr HUS/990/2017. Further details on permit and biobank decision numbers are available in the FinnGen Ethics Statement in the supplemental information.

## References

1. Leppert, B., et al A cross-disorder PRS-pheWAS of 5 major psychiatric disorders in UK Biobank. PLoS Genet 16, e1008185 (2020).

2 Mars, N., et al Polygenic and clinical risk scores and their impact on age at onset and prediction of cardiometabolic diseases and common cancers. Nat Med 26, 549–557 (2020).

3. Patel, A. P. et al. A multi-ancestry polygenic risk score improves risk prediction for coronary artery disease. Nat Med 29, 1793–1803 (2023).

4 Richardson, T. G., Harrison, S., Hemani, G. & Davey Smith, G. An atlas of polygenic risk score associations to highlight putative causal relationships across the human phenome. Elife 8, (2019).

5. Fritsche, L. G. et al. Cancer PRSweb: An Online Repository with Polygenic Risk Scores for Major Cancer Traits and Their Evaluation in Two Independent Biobanks. Am J Hum Genet 107, 815–836 (2020).

6. Ma, Y., Patil, S., Zhou, X., Mukherjee, B. & Fritsche, L. G. ExPRSweb: An online repository with polygenic risk scores for common health-related exposures. Am J Hum Genet 109 1742–1760 (2022).

7. Wand, H. et al. Improving reporting standards for polygenic scores in risk prediction studies. Nature 591, 211–219 (2021).

8 Lambert, S. A., et al The Polygenic Score Catalog as an open database for reproducibility and systematic evaluation. Nat. Genet. 53, 420–425 (2021).

9. Sun, T.-H. et al. Utility of polygenic scores across diverse diseases in a hospital cohort for predictive modeling. Nature Communications 15, 1–12 (2024).

10. Carmona, R. et al. The Spanish Polygenic Score reference distribution: a resource for personalized medicine. Eur J Hum Genet (2025) doi:10.1038/s41431-025-01850-9

11. A global reference for human genetic variation. Nature 526, 68–74 (2015).

12. Bergström, A. et al. Insights into human genetic variation and population history from 929 diverse genomes. Science 367, (2020).

13. Kurki, M. I. et al. FinnGen provides genetic insights from a well-phenotyped isolated population. Nature 613, 508–518 (2023).

14. Privé, F., Arbel, J., Aschard, H. & Vilhjálmsson, B. J. Identifying and correcting for misspecifications in GWAS summary statistics and polygenic scores. HGG Adv 3, 100136 (2022).

15. Kachuri, L. et al. Pan-cancer analysis demonstrates that integrating polygenic risk scores with modifiable risk factors improves risk prediction. Nat Commun 11, 6084 (2020).

16. Privé, F. et al. Portability of 245 polygenic scores when derived from the UK Biobank and applied to 9 ancestry groups from the same cohort. Am J Hum Genet 109, 373 (2022).

17. Bakshi, A. et al. Genomic Risk Score for Melanoma in a Prospective Study of Older Individuals J Natl Cancer Inst 113, 1379–1385 (2021).

18. Xin, J., et al Prognostic evaluation of polygenic risk score underlying pan-cancer analysis: evidence from two large-scale cohorts. EBioMedicine 89, 104454 (2023).

19. Weissbrod, O. et al. Leveraging fine-mapping and multipopulation training data to improve cross-population polygenic risk scores. Nat Genet 54, 450–458 (2022).

20. Abramowitz, S. A. et al. Evaluating Performance and Agreement of Coronary Heart Disease Polygenic Risk Scores. JAMA (2024) doi:10.1001/jama.2024.23784

21. Ding, Y. et al. Large uncertainty in individual polygenic risk score estimation impacts PRS-based risk stratification. Nat Genet 54, 30–39 (2022).

22. Lee, S. H., Goddard, M. E., Wray, N. R. & Visscher, P. M. A better coefficient of determination for genetic profile analysis. Genet Epidemiol 36, 214–224 (2012).

23. Polfus, L. M., et al Genetic discovery and risk characterization in type 2 diabetes across diverse populations. HGG Adv 2, (2021).

24. Reeve, M., et al Autoimmune hypothyroidism GWAS reveals independent autoimmune and thyroid-specific contributions and an inverse relation with cancer risk. Res Sq (2024) doi:10.21203/rs.3.rs-4626646/v

25. Campos, A. I., et al Understanding genetic risk factors for common side effects of antidepressant medications. Commun Med (Lond) 1, 45 (2021).

26. Ma, Y., Wang, M. & Zhang, Z. The association between depression and thyroid function. Front. Endocrinol 15, 1454744 (2024).

27. Hage, M. P. & Azar, S. T. The Link between Thyroid Function and Depression. J Thyroid Res 2012, 590648 (2012).

28. Dhar, A. K. & Barton, D. A. Depression and the Link with Cardiovascular Disease. Front Psychiatry 7, 33 (2016).

29. Kwapong, Y. A. et al. Association of Depression and Poor Mental Health With Cardiovascular Disease and Suboptimal Cardiovascular Health Among Young Adults in the United States. J Am Heart Assoc 12, e028332 (2023).

30. Piovani, D., Armuzzi, A. & Bonovas, S. Association of Depression With Incident Inflammatory Bowel Diseases: A Systematic Review and Meta-Analysis. Inflamm Bowel Dis 30, 573–584 (2023).

31. Ellis, C. A., et al Inflation of polygenic risk scores caused by sample overlap and relatedness: Examples of a major risk of bias. Am J Hum Genet 111, 1805–1809 (2024).

32. Hao, L. et al. Development of a clinical polygenic risk score assay and reporting workflow. Nat Med 28, 1006–1013 (2022).

33. Bakker, M. K. et al. Genetic Risk Score for Intracranial Aneurysms: Prediction of Subarachnoid Hemorrhage and Role in Clinical Heterogeneity. Stroke 54, 810–818 (2023).

34. Mars, N. et al. Systematic comparison of family history and polygenic risk across 24 common diseases. Am J Hum Genet 109, 2152–2162 (2022).

35. Michailidou, K. et al. Large-scale genotyping identifies 41 new loci associated with breast cancer risk. Nat Genet 45, 353–61, 361e1–2 (2013).

36. Haider, H., Hoehn, B., Davis, S. & Greiner, R. Effective ways to build and evaluate individual survival distributions. J. Mach. Learn. Res. 2, (2020).

37. Ge, T., Chen, C.-Y., Ni, Y., Feng, Y.-C. A. & Smoller, J. W. Polygenic prediction via Bayesian regression and continuous shrinkage priors. Nat Commun 10, 1776 (2019).

38. Privé, F., Arbel, J. & Vilhjálmsson, B. J. LDpred2: better, faster, stronger. Bioinformatics 36 5424–5431 (2021).

39. Lambert, S. A., et al Enhancing the Polygenic Score Catalog with tools for score calculation and ancestry normalization. Nat Genet 56, 1989–1994 (2024).

40. Misra, A. et al. Instability of high polygenic risk classification and mitigation by integrative scoring. Nat Commun 16, 1584 (2025).

41. Truong, B. et al. Integrative polygenic risk score improves the prediction accuracy of complex traits and diseases. Cell Genom 4, 100523 (2024).

42. Kullo, I. J. Promoting equity in polygenic risk assessment through global collaboration. Nat Genet 56, 1780–1787 (2024).

43. Wilkinson, M. D. et al. The FAIR Guiding Principles for scientific data management and stewardship. Sci Data 3, 160018 (2016).

44. World Health Organization. Accelerating Access to Genomics for Global Health: Promotion, Implementation, Collaboration, and Ethical, Legal, and Social Issues. A Report of the WHO Science Council. (World Health Organization, 2022).

45. Kurki, M. I. et al. FinnGen provides genetic insights from a well-phenotyped isolated population. Nature 613, 508–518 (2023).

46. Purcell, S. et al. PLINK: a tool set for whole-genome association and population-based linkage analyses. Am J Hum Genet 81, 559–575 (2007).

47. Marschner, I. Glm2: Fitting generalized linear models with convergence problems. R J. 3 12 (2011).

48. Privé, F., Aschard, H., Ziyatdinov, A. & Blum, M. G. B. Efficient analysis of large-scale genome-wide data with two R packages: bigstatsr and bigsnpr. Bioinformatics 34 2781–2787 (2018).

49. Reeve, M. P., et al Oral and non-oral lichen planus show genetic heterogeneity and differential risk for autoimmune disease and oral cancer. Am J Hum Genet 111, 1047–1060 (2024).

