## Supplementary Note and Figures for "PGS Browser: a public platform for personalized polygenic score interpretation"

[#] - full list of FinnGen authors is presented within Supplementary Table 13

**The conflict of interest statement is at the end of the manuscrip**

**Table of contents:**

Ethics Statement 3
PGS models processing **4**
PGS evaluation measures 4
Ancestry prediction in FinnGen cohort§ 6
Ancestry adjustment 8
External predictive models 8
References 9

*Ethics Statement*

Ethics statement and materials & methods:
Study subjects in FinnGen provided informed consent for biobank research, based on the Finnish Biobank Act. Alternatively, separate research cohorts, collected prior the Finnish Biobank Act came into effect (in September 2013) and start of FinnGen (August 2017), were collected based on study-specific consents and later transferred to the Finnish biobanks after approval by Fimea (Finnish Medicines Agency), the National Supervisory Authority for Welfare and Health. Recruitment protocols followed the biobank protocols approved by Fimea. The Coordinating Ethics Committee of the Hospital District of Helsinki and Uusimaa (HUS) statement number for the FinnGen study is Nr HUS/990/2017.
The FinnGen study is approved by Finnish Institute for Health and Welfare (permit numbers: THL/2031/6.02.00/2017, THL/1101/5.05.00/2017, THL/341/6.02.00/2018, THL/2222/6.02.00/2018, THL/283/6.02.00/2019, THL/1721/5.05.00/2019 and THL/1524/5.05.00/2020), Digital and population data service agency (permit numbers: VRK43431/2017-3, VRK/6909/2018-3, VRK/4415/2019-3), the Social Insurance Institution (permit numbers: KELA 58/522/2017, KELA 131/522/2018, KELA 70/522/2019, KELA 98/522/2019, KELA 134/522/2019, KELA 138/522/2019, KELA 2/522/2020, KELA 16/522/2020), Findata permit numbers THL/2364/14.02/2020, THL/4055/14.06.00/2020, THL/3433/14.06.00/2020, THL/4432/14.06/2020, THL/5189/14.06/2020, THL/5894/14.06.00/2020, THL/6619/14.06.00/2020, THL/209/14.06.00/2021, THL/688/14.06.00/2021, THL/1284/14.06.00/2021, THL/1965/14.06.00/2021, THL/5546/14.02.00/2020, THL/2658/14.06.00/2021, THL/4235/14.06.00/2021, Statistics Finland (permit numbers: TK-53-1041-17 and TK/143/07.03.00/2020 (earlier TK-53-90-20) TK/1735/07.03.00/2021, TK/3112/07.03.00/2021) and Finnish Registry for Kidney Diseases permission/extract from the meeting minutes on 4th July 2019. The Biobank Access Decisions for FinnGen samples and data utilized in FinnGen Data Freeze 11 include: THL Biobank BB2017_55, BB2017_111, BB2018_19, BB_2018_34, BB_2018_67, BB2018_71, BB2019_7, BB2019_8, BB2019_26, BB2020_1, BB2021_65, Finnish Red Cross Blood Service Biobank 7.12.2017, Helsinki Biobank HUS/359/2017, HUS/248/2020, HUS/430/2021 §28, §29, HUS/150/2022 §12, §13, §14, §15, §16, §17, §18, §23, §58 and §59, Auria Biobank AB17-5154 and amendment #1 (August 17 2020) and amendments BB_2021-0140, BB_2021-0156 (August 26 2021, Feb 2 2022), BB_2021-0169, BB_2021-0179, BB_2021-0161, AB20-5926 and amendment #1 (April 23 2020) and it´s modification (Sep 22 2021), BB_2022-0262, BB_2022-0256, Biobank Borealis of Northern Finland_2017_1013, 2021_5010, 2021_5018, 2021_5015, 2021_5015 Amendment, 2021_5023, 2021_5023 Amendment, 2021_5017, 2022_6001, 2022_6006 Amendment, BB22-0067, 2022_0262, Biobank of Eastern Finland 1186/2018 and amendment 22§/2020, 53§/2021, 13§/2022, 14§/2022, 15§/2022, 27§/2022, 28§/2022, 29§/2022, 33§/2022, 35§/2022, 36§/2022, 37§/2022, 39§/2022, 7§/2023, Finnish Clinical Biobank Tampere MH0004 and amendments (21.02.2020 & 06.10.2020), 8§/2021, 9§/2021, §9/2022, §10/2022, §12/2022, 13§/2022, §20/2022, §21/2022, §22/2022, §23/2022, 28§/2022, 29§/2022, 30§/2022, 31§/2022, 32§/2022, 38§/2022, 40§/2022, 42§/2022, 1§/2023, Central Finland Biobank 1-2017, BB_2021-0161, BB_2021-0169, BB_2021-0179, BB_2021-0170, BB_2022-0256, and Terveystalo Biobank STB 2018001 and amendment 25th Aug 2020, Finnish Hematological

Registry and Clinical Biobank decision 18th June 2021, Arctic biobank P0844: ARC_2021_1001.

*PGS models processing*

Most preprocessing steps were carried out using the *pgscatalog-utils* Python package (see **Web Resources**). We began by downloading 3,688 PGS Catalog models using the *download_scorefiles* function. For computational efficiency and parallel processing, all models were then merged into five separate matrices using *combine_scorefiles*. To harmonize the PGS models (GRCh38) with FinnGen genotype data (Release 11), we used the .bim file and applied the *match_variants* function, using the default matching threshold of 75%. Models that did not meet this threshold were excluded from further analysis. PGS scores were computed for each pair of matching score files (**_ALL_additive_[01].scorefile.gz.tsv*) using PLINK's *--score* function with the *cols=+scoresums,+denom* arguments. The final score for each individual was calculated as the sum of matched SNP weights divided by the number of non-missing alleles. Finally, we manually reviewed each model for potential technical inconsistencies, including misreported effect alleles, incorrect trait labels, and population mismatches between the GWAS cohort and FinnGen.

*PGS evaluation measures*

To evaluate PGS performance in this article we utilized several measures:

1) Unadjusted area under the receiver operating characteristic curve (ROC $AUC_{unadjust.}$)

ROC AUC and corresponding confidence intervals (CIs) were calculated using *AUCBoot* function from the *bigsnpr* R package [1]. The ROC AUC is defined as the probability that the predictive model will rank a randomly chosen positive instance higher than a randomly chosen negative instance. Formally:

$$ROCAUC = P(S(x_{pos}) > S(x_{neg})),$$

where $S(x)$ is the score assigned by the model to instance $x$, $x_{pos}$ and $x_{neg}$ are randomly chosen positive and negative instances, respectively. Note that sometimes ROC AUC can be less than 0.5, which means that cases are placed by PGS model lower than controls. In most of the cases this is the result of effective allele misreporting, the result of the extremely small number of cases (n<5) or drastic ancestry mismatch between target cohort and GWAS used to derive the PGS model. Note, that ROC AUC using this method was calculated for raw PGS vectors, without including any covariates.

2) Variance explained (liability log-scale)

Variance explained ($R^2$) on the logistic liability scale quantifies how much of the variance in the log-odds of the outcome is explained by the predictors. In this article we reported incremental value, calculated as a difference between *full model* value (with covariates PGS, Age, PC1-6) and *null model* (excluding PGS). In order to calculate this variance we used approach described in Lee et al [2]:

```
lrmv = glm(y~g,family = binomial(logit)) # logistic model
R2 = var(lrmv$linear.predictors)/(var(lrmv$linear.predictors)+pi^2/3)
```

3) Odds Ratio/Hazard Ratio

Odds Ratio (OR) and Hazard Ratio (HR) are a measure of association between PGS and an outcome produced by the logistic regression and cox-proportional hazard model, respectively. Both of them were calculated as an exponent of the $\beta_{PGS}$ coefficient produced by the selected models. We adjusted the relationship between standardized PGS and each endpoint for Sex, Age, PC1-6, and Genotyping Array (see **Materials and Methods, Phenome-Wide Association Study**).

4) Recall (True Positive Rate) and Specificity (True Negative Rate)

Recall (or TPR) is the proportion of actual positive cases that are correctly identified for the selected PGS threshold. Specificity (or TNR) is the proportion of actual negative cases that are correctly identified for the selected PGS threshold:

$$\mathrm{TPR} = \frac{\mathrm{TP}}{\mathrm{TP}+\mathrm{FN}} \qquad \mathrm{TNR} = \frac{\mathrm{TN}}{\mathrm{TN}+\mathrm{FP}},$$

where TP = true positives, FN = false negatives, TN = true negatives, FP = false positives.

5) Positive Predictive Value (PPV), False Discovery Rate (FDR) and Standardized Positive Predictive Value ($PPV_{std}$)

The Positive Predictive Value (PPV) is the fraction of predicted positive instances that are actually positive. Opposite to this measure is False Discovery Rate (FDR):

$$\mathrm{PPV} = \frac{\mathrm{TP}}{\mathrm{TP}+\mathrm{FP}} \qquad \mathrm{FDR} = \frac{\mathrm{FP}}{\mathrm{TP}+\mathrm{FP}} = 1-\mathrm{PPV}$$

We used a standardized PPV ($PPV_{std}$) [3] to make PPV comparable between different disease prevalence. During this procedure specificity and sensitivity get fixed and reference prevalence gets selected. We choose a prevalence of 5% as a reference value, since it represents around the median value of prevalence of diseases tested. Adjustment procedure goes like that:

$$PPV_{std} = \frac{\text{Sensitivity} \times \text{Prevalence}}{(\text{Sensitivity} \times \text{Prevalence}) + ((1-\text{Specificity}) \times (1-\text{Prevalence}))}$$

6) Absolute/Relative Risk

Absolute risk was estimated via Bayes' theorem as the posterior probability of having disease given a specific PGS threshold:

$$P(\text{Case} \mid \text{PGS}) = \frac{P(\text{PGS} \mid \text{Case}) \times P(\text{Case})}{P(\text{PGS} \mid \text{Case}) \times P(\text{Case}) + P(\text{PGS} \mid \text{Control}) \times P(\text{Control})}$$

Relative Risk was defined as the probability of being a case for the selected PGS threshold divided by the probability of being the case at 50th PGS percentile.

7) Time-dependent area under the ROC curve ($_{td}$ROC AUC)

$_{td}$ROC AUC is a time-dependent extension of ROC AUC calculated over preselected time-points and averaged. It was calculated using *cumulative_dynamic_auc* function from *scikit-survival* (v.0.23.0) Python package. We used this measure to evaluate the performance of survival models. Selected time-points are mentioned in corresponding figure captions. In order to estimate the confidence intervals, we bootstrapped the testing set, calculated average $_{td}$ROC AUC across different time points and, in the end, derived CIs from the sample of 100 average $_{td}$ROC AUCs. It is important to note that this measure just tells how well cases are separated from controls and does not consider how well probabilities and produced survival functions are calibrated.

8) Integrated Brier Score (IBS)

IBS was calculated using *integrated_brier_score* from *scikit-survival* (v.0.23.0) Python package. It evaluates both discrimination and calibration of survival models, lower values depicting better performing models. For details, please see description in the *scikit-survival* documentation.Time points matched those used in $_{td}$ROC AUC calculations.

9) D-calibration

To assess the calibration of predicted survival functions, we employed D-calibration [4]. P-value for D-calibration was calculated using *SurvivalEval* (v.0.3.0) python package. Corresponding p-values reflect the goodness-of-fit between predicted and observed event probabilities, thus p-value>0.05 represents a well-calibrated model.

*Ancestry prediction in FinnGen cohort§*

To model continental population structure, we first selected 51,137 HapMap3 variants shared between the 1000 Genomes Project whole-genome sequencing (WGS) data and the FinnGen genotyping array. Restricting principal components analysis (PCA) to these variants we tried to exclude the batch effect introduced by the imputation and at the same time keep variants informative for the ancestry prediction in FinnGen samples. Using PLINK's *-pca* we derived six principal components and projected all 473,681 FinnGen samples into this 1000G PC space (**Fig. 5a**). Each PC-score was standardized by dividing by the square root of its eigenvalue and scaling by –2. Further, we used these PCs to infer ancestry for non-Finnish samples.

To infer ancestry for non-Finnish samples, we trained an XGBoost classifier (*xgboost*, v3.0.2) on 1000 Genomes data (90% training, 10% testing). We optimized the following hyperparameters using grid search (cv=10) and one-vs-rest ROC AUC as a scoring function: *n_estimators*, *max_depth*, *learning_rate*, *subsample*, *colsample_bytree*, and *reg_lambda*. The final model achieved a weighted F1-score of 0.99 on the test set.

Using the tuned XGBoost classifier, we assigned continental ancestries to 19,947 non-Finnish FinnGen participants: 15,101 non-Finnish Europeans, 1,620 Admixed Americans, 690 East Asians, 433 Africans, and 352 South Asians. Individuals whose highest ancestry probability fell below 65% (n = 1,751) were labeled as “Other.”

*Disease risk prediction models*

To identify diseases that benefit from integrating multiple PGSs for diagnosis prediction or future incidence risk, we developed predictive models for both disease status classification and age-of-onset prediction. These models were designed to assess whether incorporating multiple polygenic scores improves predictive performance beyond the use of a single best-performing PGS. All models were trained and tested on 453,734 Finnish ancestry samples with complete covariate data. To make sure that there is no sample overlap between GWASs used for PGS models development and FinnGen samples, for further experiments we utilized solely top 6 publications, covered almost the half of all available models (n=1,514), which reliably lack any FinnGen sample overlap, naming: Tanigawa Y et al. (PGP000244), Privé F et al. (PGP000263), Weissbrod O et al. (PGP000332), Liu N et al. (PGP000464), Ding Y et al. (PGP000457) and Sinnott-Armstrong N et al. (PGP000128) [5–10].

For each disease endpoint, we trained three distinct models with different sets of covariates. First, *null model* - included sex, age, and the first six genetic principal components (PCs) as predictors. For binary classification (disease status), age was defined as the age at the end of follow-up. For age-of-onset prediction, age was defined as baseline age at recruitment. Second, *best model* - extended the *null model* by incorporating the single best-performing PGS, determined based on the highest ROC AUC in the training set. This selection process ensured that no data leakage occurred from the test set. Third, *multi model* - included the *null model* covariates along with an optimal combination of 1,514 PGSs, selected through elastic net regularization. This approach was applied to both binary classification and age-of-onset prediction. In most of the cases, the majority of PGSs after regularization had weight of zero, thus in the end only a small subset of PGSs were taken for model training.

For diagnosis prediction, we trained a logistic regression model with elastic net regularization (penalty='*elasticnet*'), implemented in the *scikit-learn* (v.1.5.2) Python package. To ensure proper model fitting and comparability across features, we applied standardization to all predictors before model training. Hyperparameter tuning was performed using a randomized search strategy, using ROC AUC as the scoring metric. We optimized: the inverse regularization strength (*C*) and L1 regularization ratio (*l1_ratio*). A total of 200 iterations with 3-fold cross-validation were conducted. The best-performing model from this search was then applied to the test set. Corresponding results were reported in the **Supplementary Table 7**.

For age-of-onset prediction, we trained a Cox proportional hazards model with elastic net regularization (CoxNet), implemented in the *scikit-survival* (v.0.23.0) Python package. As in the classification models, all features were standardized before training. Model selection and hyperparameter tuning followed the same randomized search approach, ensuring robust performance evaluation. The only difference was that we excluded patients with baseline age less than age of the disease onset or age of follow up, thus sample sizes between diagnosis prediction and age-of-onset differed and for each disease there was a unique number of patients satisfying this filtering procedure. We used years of follow up as a time scale.

Further, we systematically compared *null*, *best*, and *multi models*. We aimed to quantify the added predictive value, comparing, first, single best PGS with null model and, second, multiple PGSs with the null model. For binary prediction we utilized ROC AUC, for survival model we checked time-dependent ROC AUC, Brier Score and D-calibration measures (see **Supplementary Note, PGS evaluation measures**). Corresponding results were reported in the **Supplementary Table 8**.

Feature importances, defined as an absolute value of the elastic net model coefficients, were extracted from Logistic Regression and CoxNet *multi models* to define optimal combination of PGSs. All scores with weight of zero were excluded. Corresponding weights were reported in the **Supplementary Table 9,10**.

*Ancestry adjustment*

Raw PGS values for individual patients can be biased by underlying population structure, so we implemented the adjustment method described in Hao et al.[11] to remove ancestry-driven variance. First, we regressed each *raw* PGS on genetic principal components (PCs) (see **Supplementary Materials, Ancestry prediction in FinnGen cohort**) to predict the ancestry-driven component (*predicted* PGS). For each of the 3,168 PGSs we used a 6-fold cross-validation approach to derive this component for every FinnGen participant. Next, we subtracted *predicted* PGS from the *raw* score. The resulting *adjusted* PGS retains the genetic signal while minimizing the variance attributed solely to population structure:

$$PGS_{adjusted} = PGS_{raw} - PGS_{predicted}$$

Notably, all *predicted* PGSs exhibited a statistically significant correlation with their respective *raw* PGSs, confirming the influence of population structure on all of the PGSs (**Figure 5c**). Expectedly, *predicted* PGSs were completely explained by the PCs and *adjusted* PGSs were completely decorrelated with PCs (**Supplementary Figure S10**).

*External predictive models*

As a part of PGS Browser functionality we provide 22 best-PGS CoxNet models which can be utilized by the external user. As an input they take biological sex, current age of the patient and *adjusted* PGS percentile, which can be calculated locally by the user. Thus, no PC information needed or individual-level data transmitted to the browser. All of these models were perfectly calibrated (D-calibration p-value > 0.99), had time-dependent AUC (1-10 year) > 0.65 and increment in AUC from inclusion of PGS at least of 1% (**Supplementary Fig. 14**). They were finetuned using the same procedures described previously (see **Supplementary Note, Disease risk prediction models**).

To automate percentile calculation for an ancestry-adjusted PGS in any individual, we developed *pgsb-cli* (see Documentation page at www.pgs.nchigm.org) - a Docker-based command-line tool that runs entirely on the user's machine: it takes as an input genotype data (vcf or plink format) and a PGS model downloaded from the PGS Browser, matches variants, projects the sample onto 1000 Genomes principal components, computes the *predicted* and *adjusted* PGSs, and outputs standardized scores and percentiles, all without transmitting any individual-level data outside the local container.

**References**


1. Privé, F., Aschard, H., Ziyatdinov, A. & Blum, M. G. B. Efficient analysis of large-scale genome-wide data with two R packages: bigstatsr and bigsnpr. *Bioinformatics* **34**, 2781–2787 (2018).
2. Lee, S. H., Goddard, M. E., Wray, N. R. & Visscher, P. M. A better coefficient of determination for genetic profile analysis. *Genet Epidemiol* **36**, 214–224 (2012).
3. Heston, T. F. Standardizing predictive values in diagnostic imaging research. *J Magn Reson Imaging* **33**, 505; author reply 506–7 (2011).
4. Haider, H., Hoehn, B., Davis, S. & Greiner, R. Effective ways to build and evaluate individual survival distributions. *J. Mach. Learn. Res.* **21**, (2020).
5. Tanigawa, Y. *et al.* Significant sparse polygenic risk scores across 813 traits in UK Biobank. *PLoS Genet* **18**, e1010105 (2022).
6. Privé, F. *et al.* Portability of 245 polygenic scores when derived from the UK Biobank and applied to 9 ancestry groups from the same cohort. *Am J Hum Genet* **109**, 373 (2022).
7. Weissbrod, O. *et al.* Leveraging fine-mapping and multipopulation training data to improve cross-population polygenic risk scores. *Nat Genet* **54**, 450–458 (2022).
8. Liu, N. *et al.* Cross-ancestry genome-wide association meta-analyses of hippocampal and subfield volumes. *Nat Genet* **55**, 1126–1137 (2023).
9. Ding, Y. *et al.* Polygenic scoring accuracy varies across the genetic ancestry continuum. *Nature* **618**, 774–781 (2023).
10. Sinnott-Armstrong, N. *et al.* Genetics of 35 blood and urine biomarkers in the UK Biobank. *Nat Genet* **53**, 185–194 (2021).
11. Hao, L. *et al.* Development of a clinical polygenic risk score assay and reporting workflow. *Nat Med* **28**, 1006–1013 (2022).
12. A global reference for human genetic variation. *Nature* **526**, 68–74 (2015).

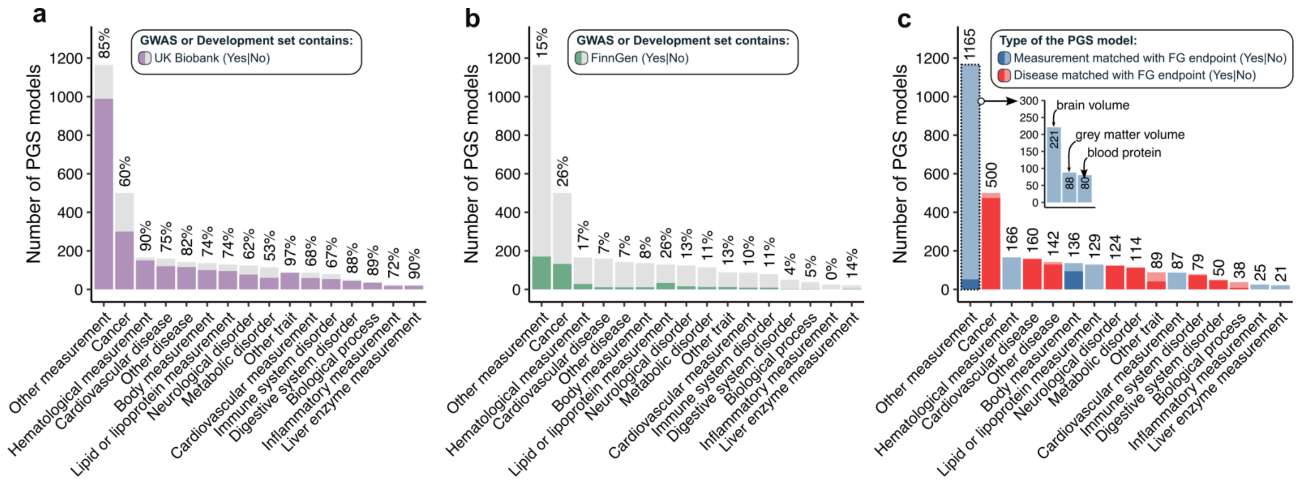


**Supplementary Figure S1. PGS Models from the PGS Catalog included in the analysis.** (a) Number of PGS models in the PGS Catalog that used the UK Biobank as a source of effect sizes or as a development set during model creation. (b) Number of PGS models in the PGS Catalog that used the FinnGen cohort as a source of effect sizes or as a development set during model creation. (c) Number of PGS models across different trait categories, as classified in the PGS Catalog. Colors indicate the type of trait: blue represents measurements, and red represents binary disease/trait status. Shaded areas (for both measurements and diseases) indicate models matched to FinnGen data, while transparent areas indicate models lacking corresponding endpoints in the FinnGen database.

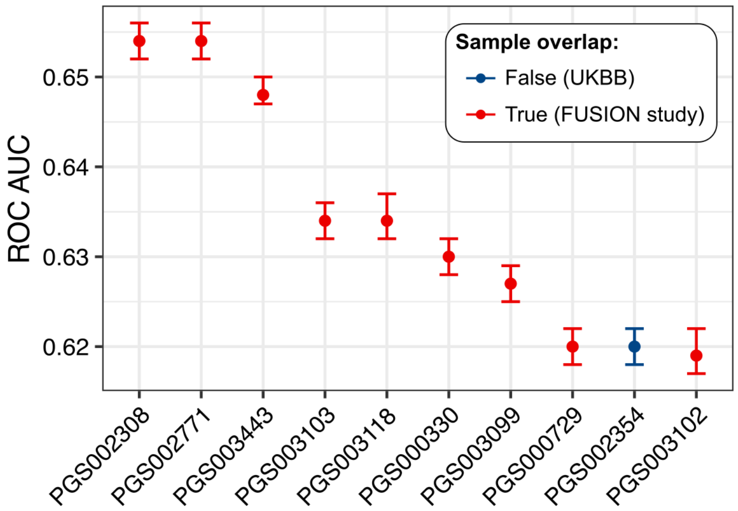


**Supplementary Figure S2. Top 10 polygenic scores for type 2 diabetes.** Colors indicate sample overlap with the FinnGen cohort. Red dots - ROC AUCs from scores derived using GWAS that include the Finland–United States Investigation of NIDDM Genetics (FUSION) cohort, a legacy cohort of FinnGen. Blue dots - the sole score derived from UK Biobank summary statistics. Of the scores shown, only PGS003443 and PGS000729 explicitly state that their GWAS included the DIAGRAM consortium. Even then, confirming that the FUSION cohort participated in score development requires a manual check of DIAGRAM's constituent sub-cohorts.

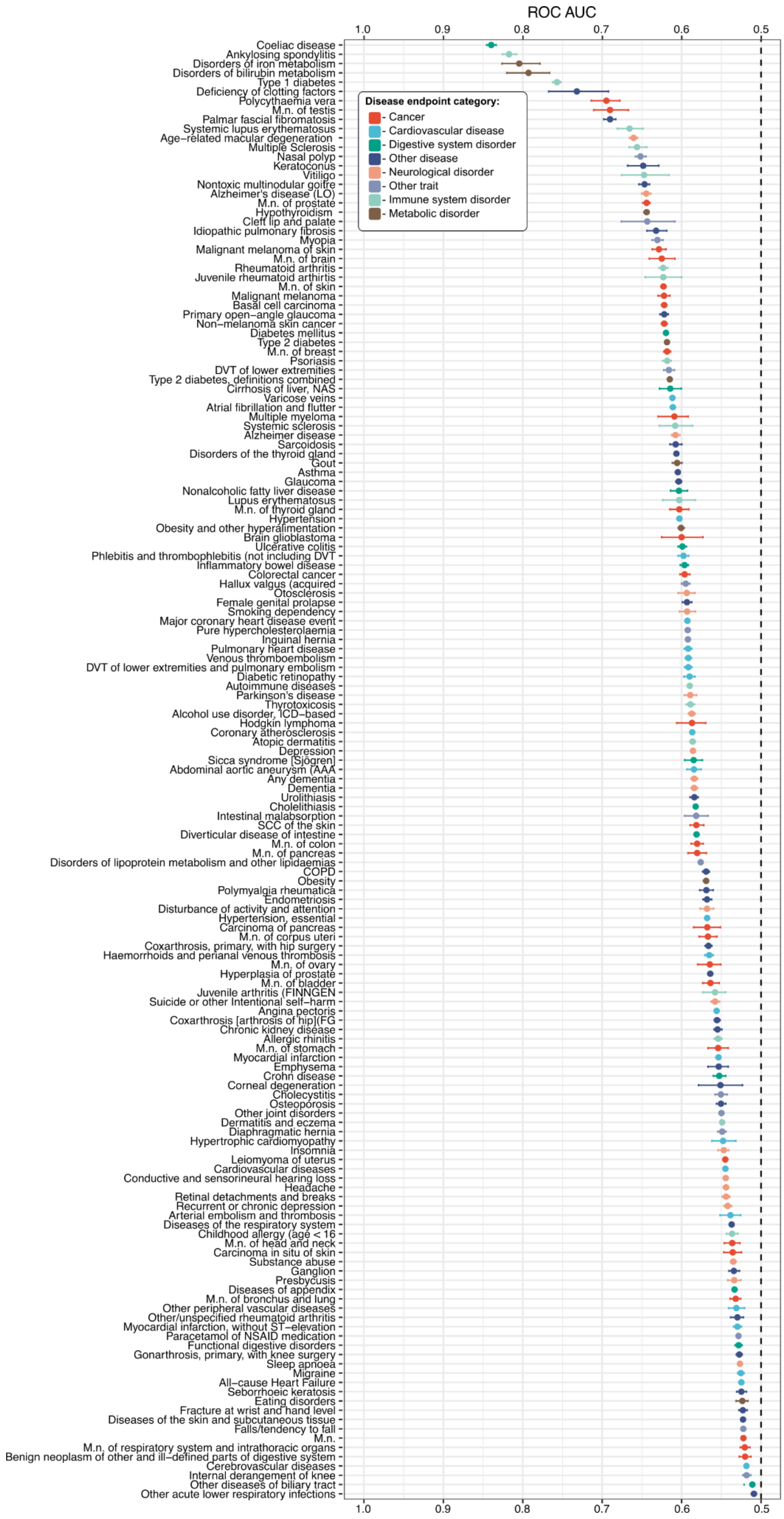


**Supplementary Figure S3.** Area under the receiver operating characteristic curve (ROC AUC) for 157 FinnGen disease endpoints. Dashed line depict ROC AUC equals 0.5. Color depicts broad disease category derived from PGS Catalog classification. FinnGen disease endpoints were additoanlly preprocessed to depict true control distributions - all samples removed from controls in original FinnGen endpoint were reintroduced and treated as controls (see https://risteys.finngen.fi/).

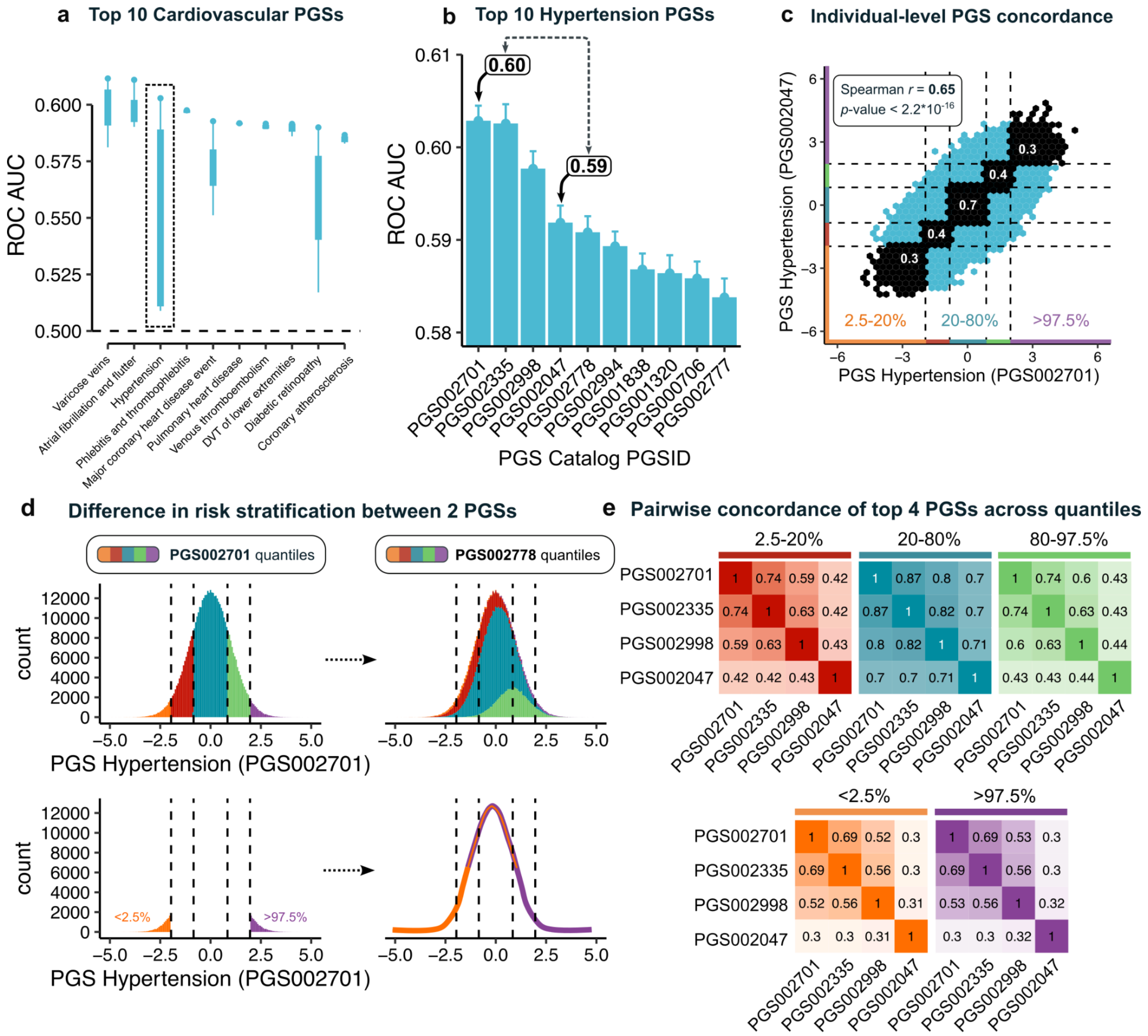


**Supplementary Figure S4.** Individual-level concordance between the top four Hypertension PGSs. (a) Top ten cardiovascular system PGSs. The dashed box highlights the distribution of ROC AUCs for hypertension PGSs. (b) Top ten hypertension PGSs. The first and fourth PGSs are compared. (c) Individual-level PGS concordance between the first and fourth PGSs from b. Colors indicate PGS distribution quantiles: orange (<2.5%), red (2.5–20%), turquoise (20–80%), green (80–97.5%), and violet (>97.5%). Numbers represent concordance between identical quantiles of the two PGSs. (d) Differences in risk stratification between the two PGSs, with colors depicting quantiles. (e) Pairwise concordance of the top four hypertension PGSs across quantiles.

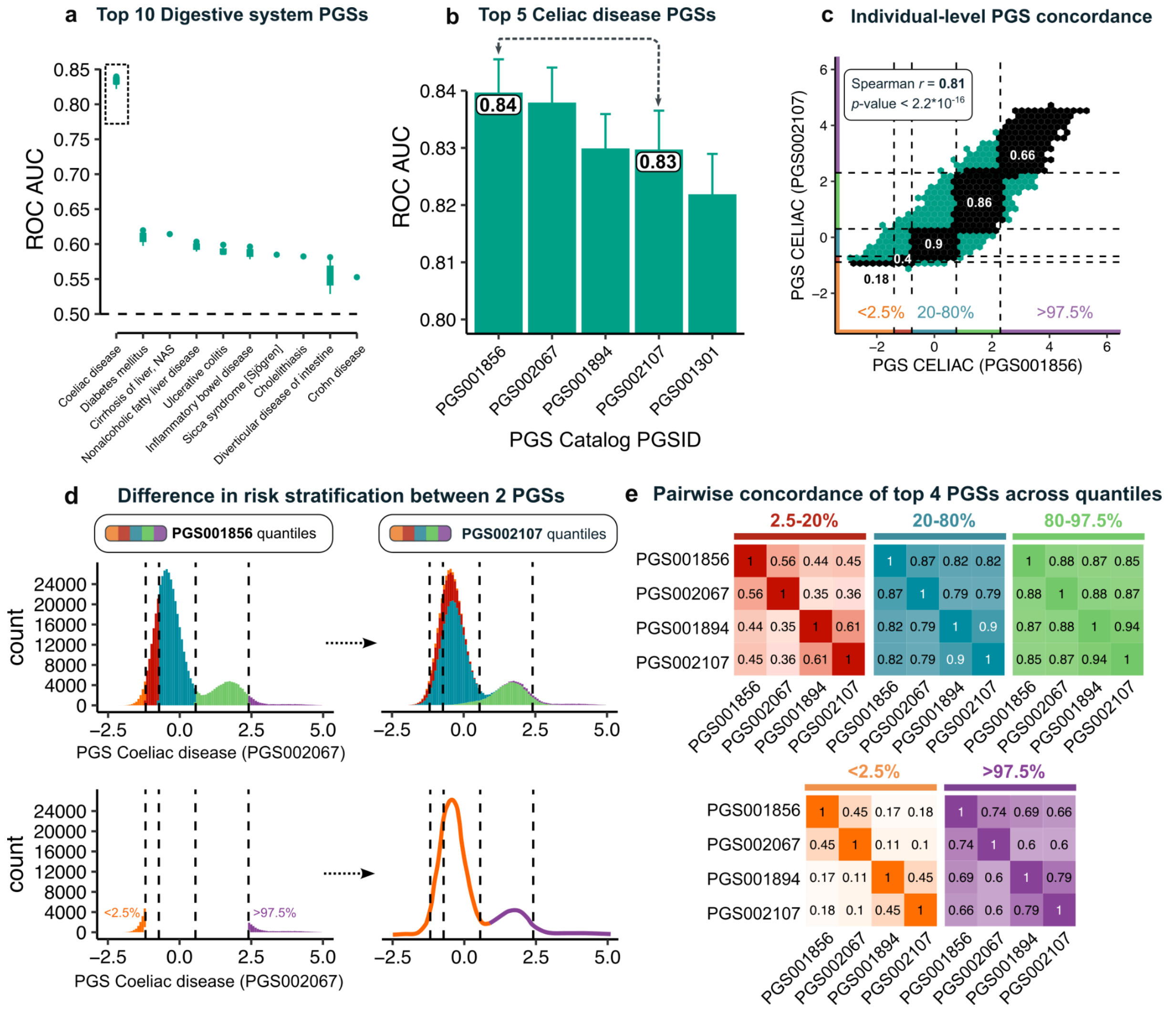


**Supplementary Figure S5.** Individual-level concordance between the top four Celiac disease PGSs. (a) Top ten digestive system PGSs. The dashed box highlights the distribution of ROC AUCs for Celiac disease PGSs. (b) Top five Celiac disease PGSs, no statistically significant differences among the top four were found. The first and fourth PGSs are compared. (c) Individual-level PGS concordance between the first and fourth PGSs from b. Colors indicate PGS distribution quantiles: orange (<2.5%), red (2.5–20%), turquoise (20–80%), green (80–97.5%), and violet (>97.5%). Numbers represent concordance between identical quantiles of the two PGSs. (d) Differences in risk stratification between the two PGSs, with colors depicting quantiles. (e) Pairwise concordance of the top four Celiac disease PGSs across quantiles.

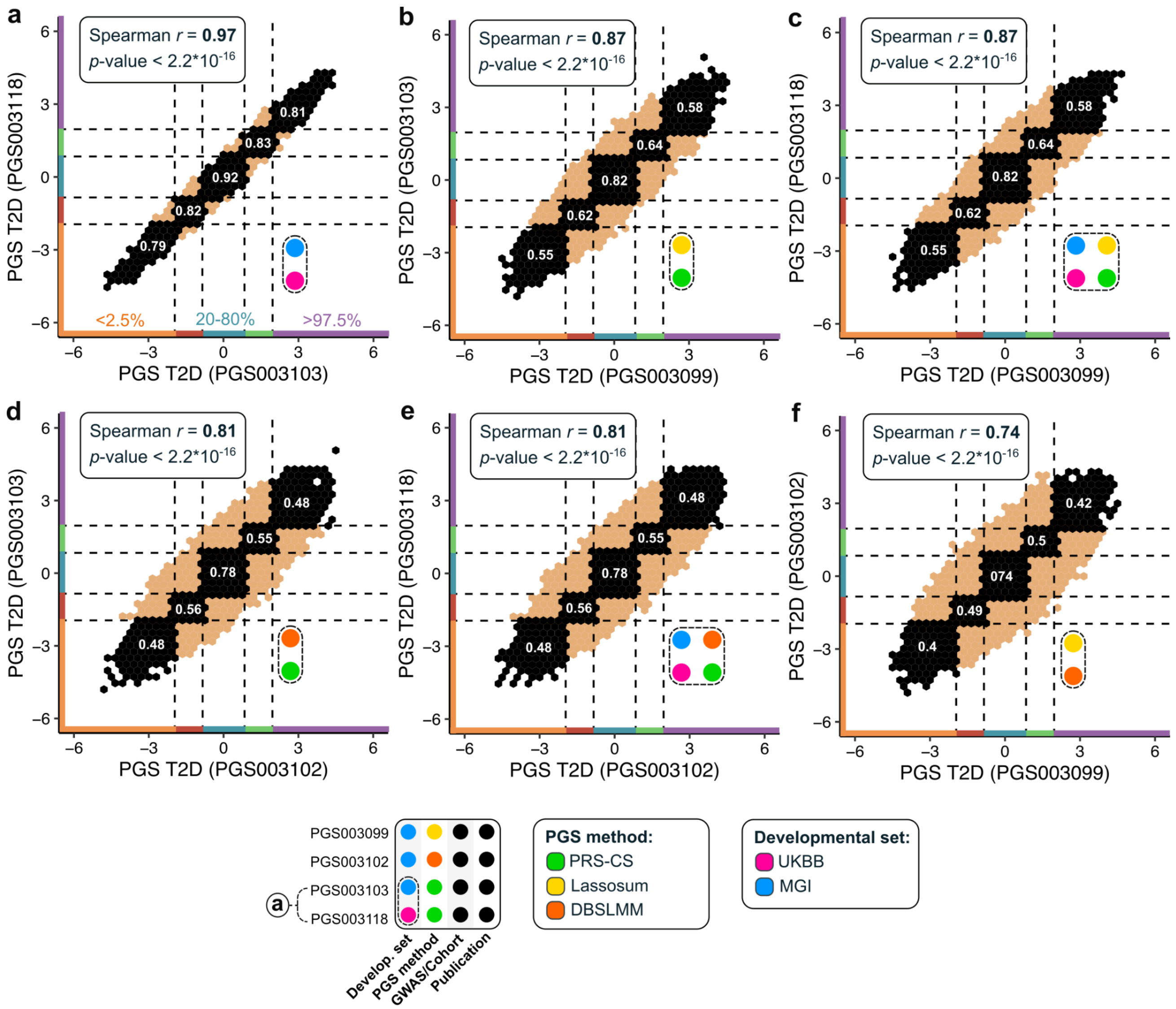


**Supplementary Figure S6.** Individual-level concordance among the top four type 2 diabetes PGSs derived from the same GWAS. The legend lists the different methodological combinations used to build each score. Even when based on identical GWAS data, these methodological choices create substantial discordance in risk stratification. White numbers in overlapping quantile segments denote the proportion of individuals shared between those quantiles.

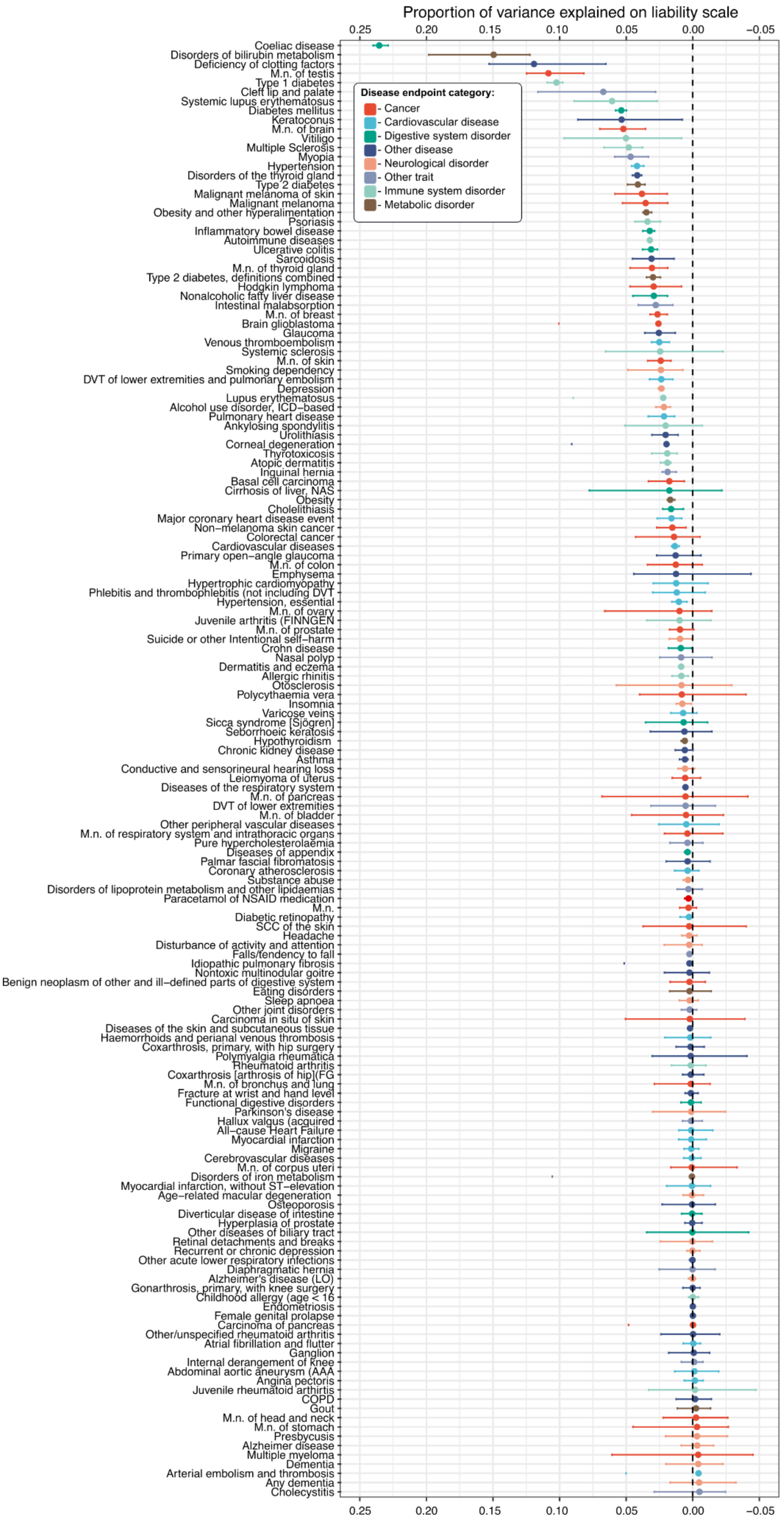


**Supplementary Figure S7.** Proportion of variance explained on logistic liability scale for 157 FinnGen disease endpoints. Dashed line depict varaince explained equals 0. Color depicts broad disease category derived from PGS Catalog classification. FinnGen disease endpoints were additoanlly preprocessed to depict true control distributions - all samples removed from controls in original FinnGen endpoint were reintroduced and treated as controls (see https://risteys.finngen.fi/).

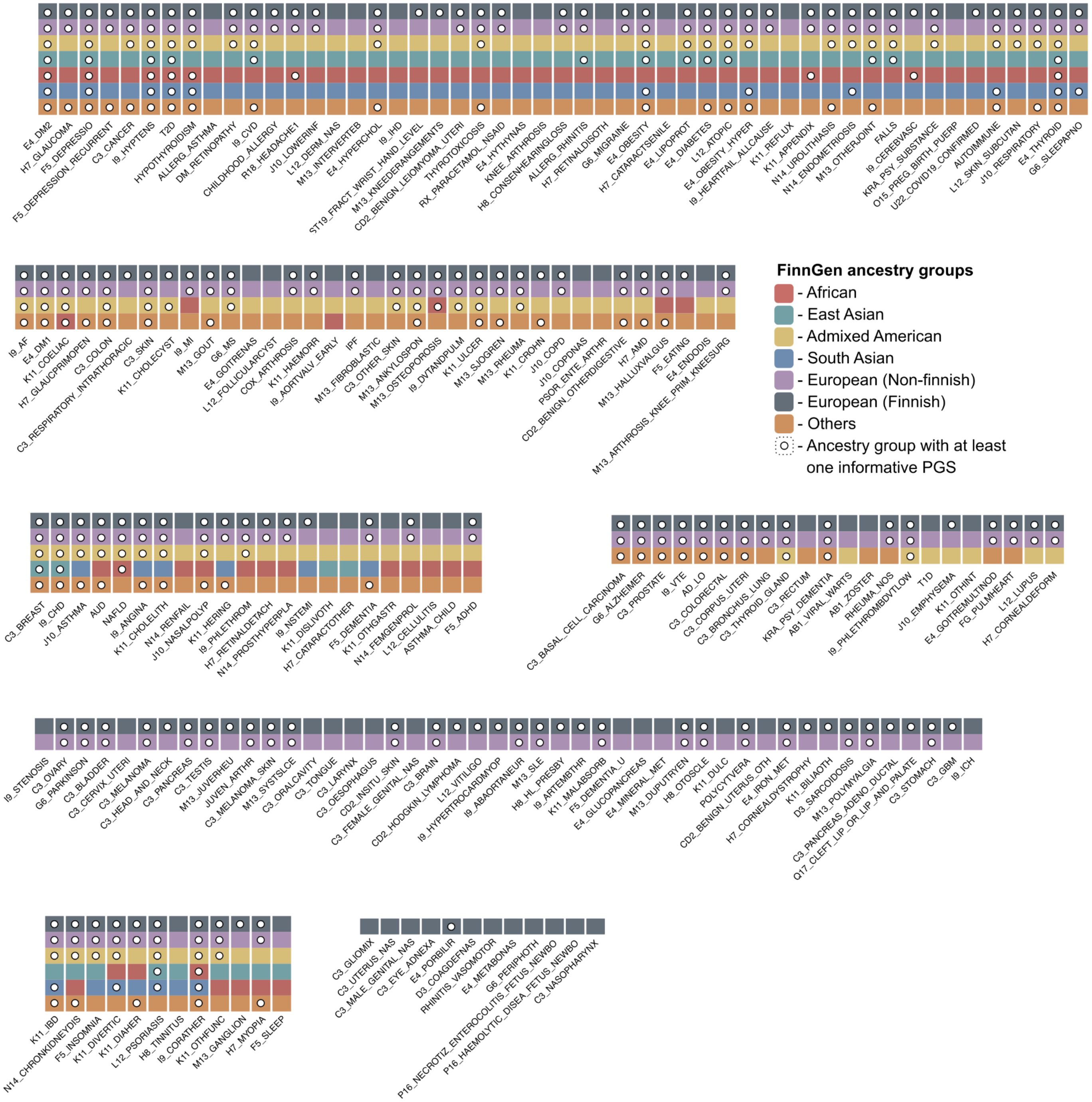


**Supplementary Figure S8.** Informative PGS presence across major ancestry groups. Coloured segments depict the ancestries analysed; white dot indicates that at least one informative PGS exists for a given endpoint–ancestry pair. Endpoints are grouped by how many ancestries meet the minimum threshold of eight cases.

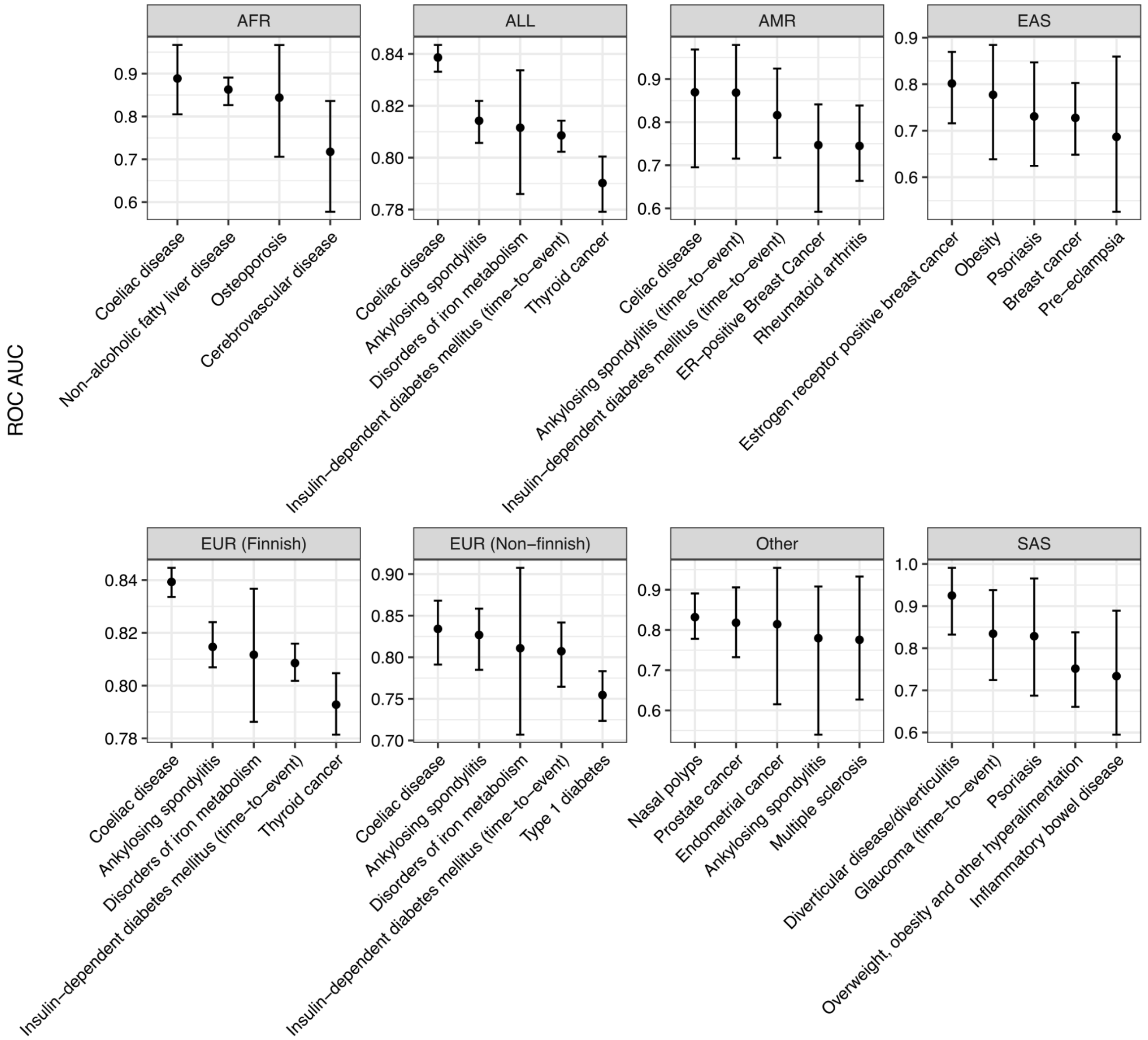


**Supplementary Figure S9.** Top five polygenic score models by ROC AUC in each ancestry group.

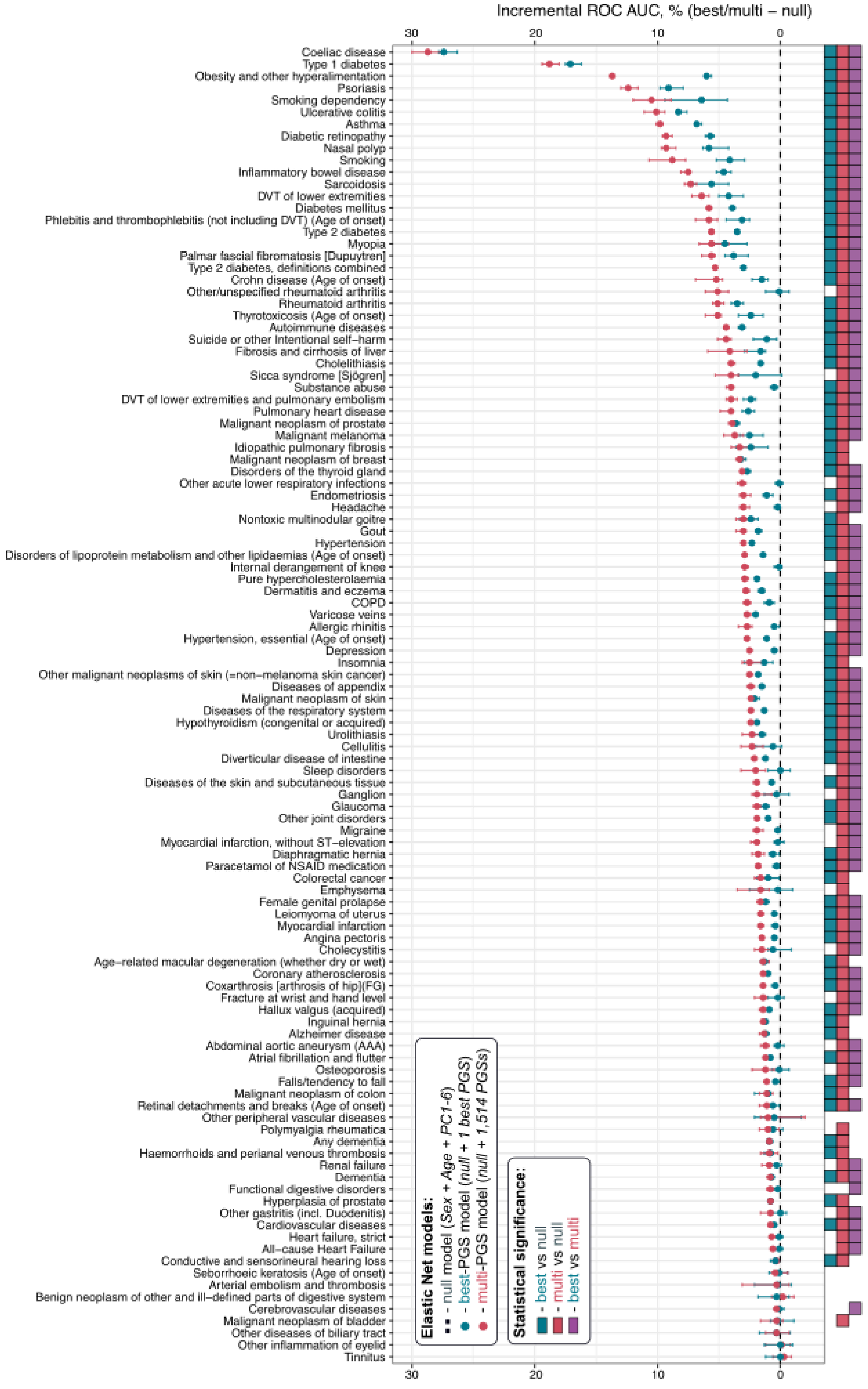


**Supplementary Figure S10**. Incremental area under the receiver operating characteristic curve (ROC AUC) between three elastic net models. "null" model included Sex, Age at the end of follow-up and first six principal components as predictors. Its incremental ROC AUC values are all zeros and were took as a baseline for each phenotype (dashed line). "best"-PGS model included predictors from "null" model and single best polygenic score (PGS) for selected phenotype (turquoise color). "multi"-PGS model included predictors from "null" model and 1,514 PGSs (red color). Colored squares depict statistical significance between three pairs of comparisons. Turquoise square - "best" model and "null" model, red - "multi" model and "null" model, violet - "best" model and "multi" model. Presence of square on the right side for selected phenotype means statistical significance in terms of ROC AUC difference on testing set.

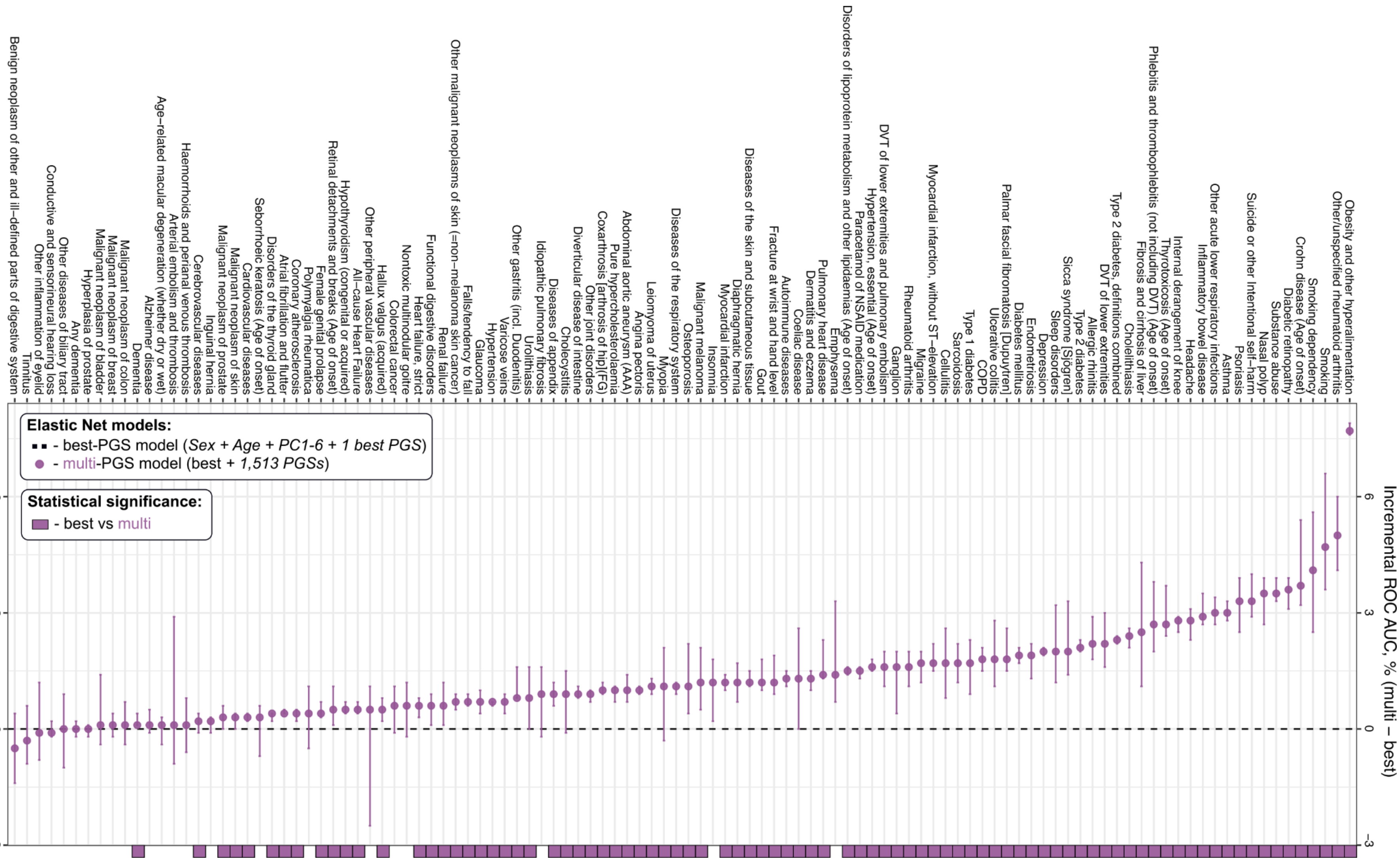


**Supplementary Figure S11**. Incremental area under the receiver operating characteristic curve (ROC AUC) between three elastic net models. "best" model included as predictors Sex, Age at the end of follow up, first six principal components and single best polygenic score (PGS) for selected phenotype. Its incremental ROC AUC values are all zeros and were took as a baseline for each phenotype (dashed line). "multi"-PGS model included predictors from "best" model and and additional 1,513 PGSs (violet color). Colored squares depict statistical significance between pair of comparison - "best" model and "multi" model. Presence of square on the right side for selected phenotype means statistical significance in terms of ROC AUC difference on testing set.

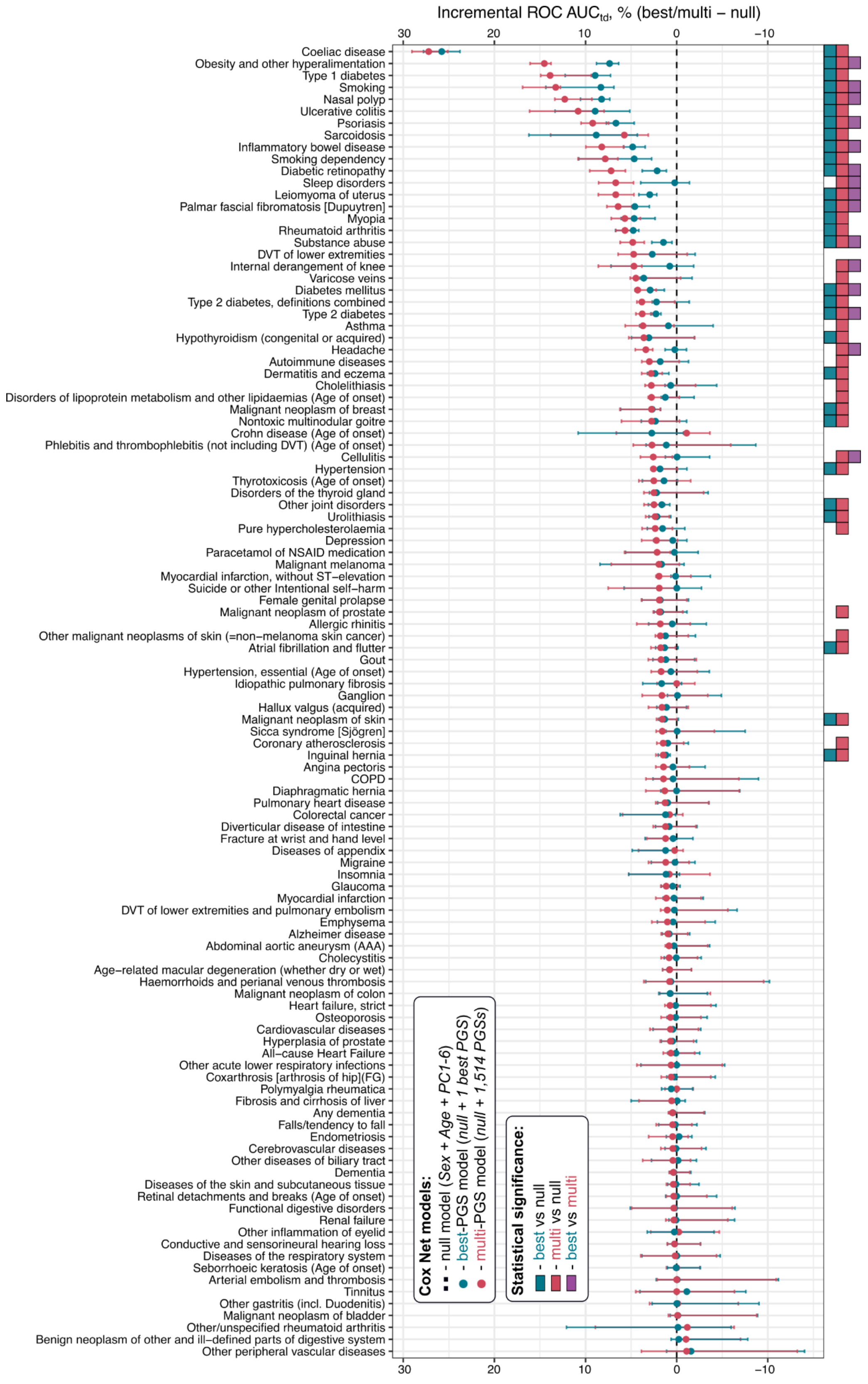


**Supplementary Figure S12**. Incremental time-dependent (td) (1-10 years) area under the receiver operating characteristic curve (ROC AUC) between three Cox's proportional hazard's models with elastic net penalty (Cox Net). "null" model included Sex, Baseline age and first six principal components as predictors. Its incremental time-dependent ROC AUC values are all zeros and were took as a baseline for each phenotype (dashed line). "best"-PGS model included predictors from "null" model and single best polygenic score (PGS) for selected phenotype (turquoise color). "multi"-PGS model included predictors from "null" model and 1,514 PGSs (red color). Colored squares depict statistical significance between three pairs of comparisons. Turquoise square - "best" model and "null" model, red - "multi" model and "null" model, violet - "best" model and "multi" model. Presence of square on the right side for selected phenotype means statistical significance in terms of time-dependent ROC AUC difference on testing set.

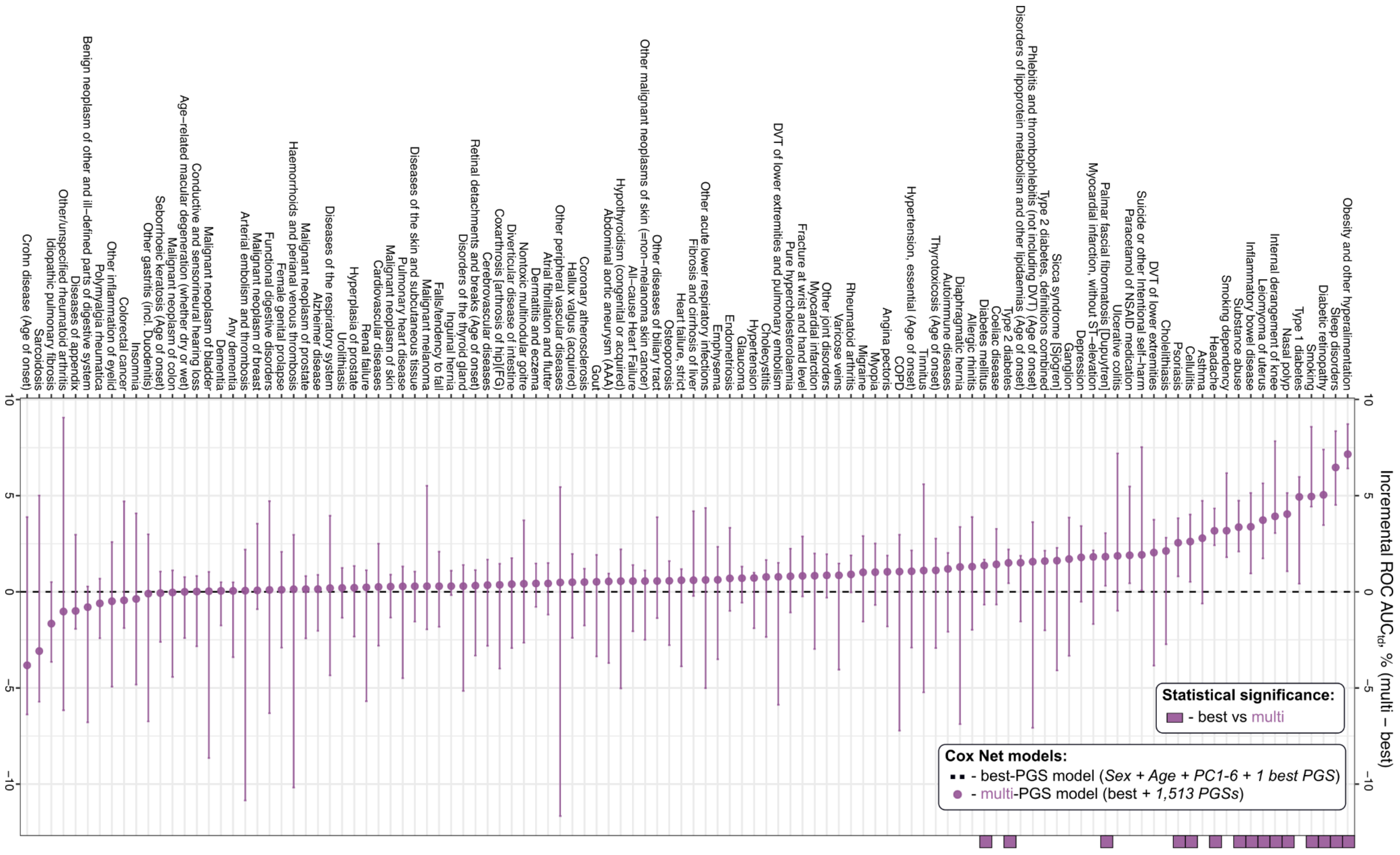


**Supplementary Figure S13.** Incremental time-dependent (1-10 years) area under the receiver operating characteristic curve (ROC AUC) between three Cox's proportional hazard's models with elastic net penalty (Cox Net). "best" model included as predictors Sex, Baseline Age, first six principal components and single best polygenic score (PGS) for selected phenotype. Its incremental ROC AUC (td) values are all zeros and were took as a baseline for each phenotype (dashed line). "multi"-PGS model included predictors from "best" model and and additional 1,513 PGSs (violet color). Colored squares depict statistical significance between pair of comparison - "best" model and "multi" model. Presence of square on the right side for selected phenotype means statistical significance in terms of ROC AUC difference on testing set.

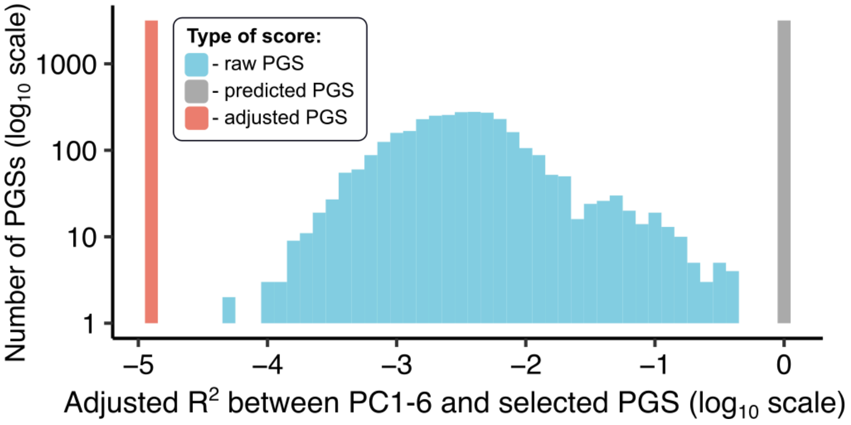


**Supplementary Figure S14.** Lack of correlation between adjusted PGS and PCs. Both X and Y-axis are in log10 scale. Blue - raw PGS, grey - predicted PGS, red - adjusted PGS.

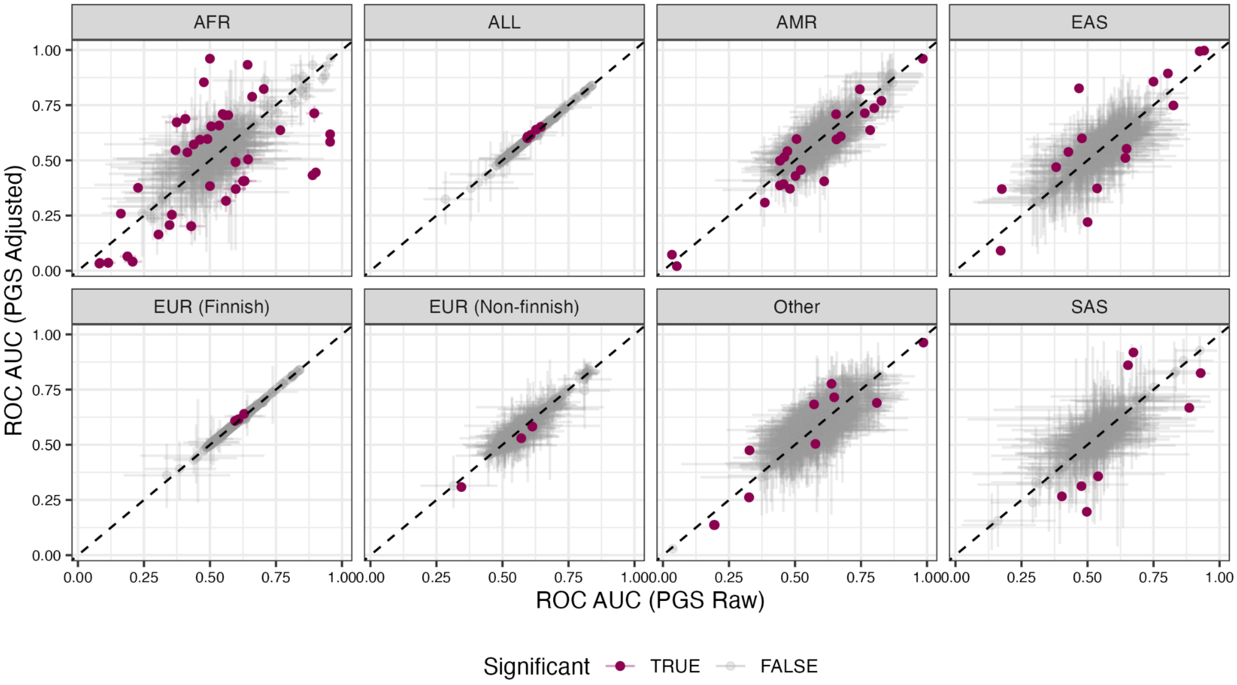


**Supplementary Figure S15** Comparison of ROC AUC values for raw and ancestry-adjusted PGSs across major ancestry groups. Points above the diagonal indicate higher accuracy after adjustment, and points below indicate a decrease. Purple color highlights scores showed significant difference in ROC AUCs after adjustment. Groups: Finnish Europeans (EUR Finnish), non-Finnish Europeans (EUR Non-Finnish), combined cohort (ALL), South Asians (SAS), “Other” group, East Asians (EAS), Admixed Americans (AMR), Africans (AFR).

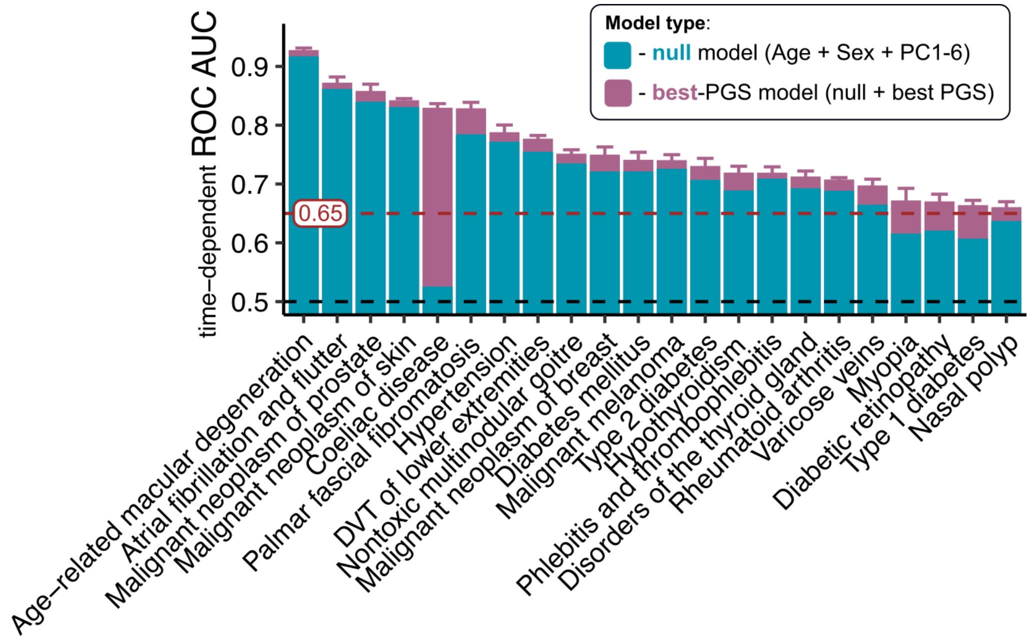


**Supplementary Figure S16** Time-dependent ROC AUCs for the 22 externally released CoxNet models.
